# Examining Age-Dependent Patterns in Academic Bullying Behaviors

**DOI:** 10.1101/2024.09.24.24314297

**Authors:** Sherry E. Moss, Morteza Mahmoudi

## Abstract

Academic bullying is a pervasive and longstanding issue that negatively affects all fields of science. While influential factors such as scientific discipline, sex, and ethnicity have been investigated, the impact of perpetrators’ age on the contextual behaviors of academic bullying has not been thoroughly studied. Our cross-sectional global survey of academic bullying, involving 2,390 participants, provides evidence that age significantly influences the contextual behaviors of academic bullying. Our results revealed that the youngest perpetrators (aged 25-35) exhibit significantly less abusive behavior compared to other age categories (36 and older). Additionally, the lowest percentage of abusive behavior was observed in the youngest group, while the highest was found in the older groups (aged 56-65). Our comprehensive analysis on the potential influence of the perpetrator’s academic position, ethnicity, or sex on their age group revealed no significant impact. These findings have strong implications for the development of more customized academic bullying training, policy development, and monitoring strategies, ultimately benefiting the scientific community at large in many aspects including evolution of science and improving the mental health of the academic workforce.

## Introduction

Academic bullying has garnered significant interest in recent years, with scientific communities across various disciplines highlighting the importance of addressing this pervasive and longstanding issue.^1-3^ Researchers have explored its negative effects not only on the immediate targets but also on the wider community and the progression of scientific research.^4^ Various influential factors, such as gender and ethnicity of both targets and perpetrators, have been investigated to better understand the root causes of academic bullying and identify vulnerable populations and likely perpetrators.^5,6^ However, the impact of perpetrators’ age on the contextual behaviors of academic bullying has not been thoroughly examined or systematically studied.

Age is a significant factor influencing a wide range of prosocial and socioemotional behaviors and actions.^7^ Emerging evidence suggests that older adults tend to be more prosocial than younger adults, as demonstrated by their increased propensity to learn about rewards for others and engage in effortful actions for the benefit of others.^8,9^ Theoretical accounts of lifespan development, such as socioemotional selectivity theory, propose that this increase in prosocial behavior is driven by age-related shifts in goals and priorities.^9,10^ As individuals age, their motivation for socially and emotionally meaningful behaviors grows, driven by a focus on immediate emotional satisfaction and meaningful social interactions. This theory suggests that older adults prioritize positive emotional experiences and meaningful connections, which in turn enhances their prosocial behaviors.

Despite extensive research on various factors contributing to academic bullying (including our own results^5,11^), the specific influence of perpetrators’ age on their bullying behaviors has not been thoroughly investigated.^6,12-16^ Understanding whether aging has similar socioemotional effects on academic bullying and harassment as it does on general prosocial behavior is crucial. This paper aims to investigate the hypothesis that aging has comparable socioemotional effects on academic bullying and harassment, potentially influencing the nature and extent of bullying behaviors exhibited by older versus younger perpetrators. Exploring the relationship between age and academic bullying behaviors can provide valuable insights into the dynamics of bullying in academic settings. It can help identify whether older individuals are more likely to engage in or refrain from bullying behaviors due to their socioemotional development. Furthermore, this research can inform the development of age-specific interventions and policies aimed at mitigating academic bullying, thereby fostering a healthier and more inclusive academic environment.

## Results and discussion

To investigate the potential role of perpetrators’ age in academic bullying, we analyzed data from our cross-sectional global survey on the topic. Full details about IRB approval [Wake Forest University (IRB00023594) and Michigan State University (STUDY00003215)], consent, and the declaration of informed consent for data usage are provided in the survey documentation (see our earlier publication^5^ for details). Data were collected from 2,390 individuals, solicited through various channels including advertorial pieces and third-party emails in Science^17^ and Nature magazines, as well as online panel advertisements and third-party emails from the American Chemical Society.

Because targets are unlikely to know the exact chronological age of their perpetrators, we asked them to estimate their perpetrators’ age group using the following 5 categories: 25-35, 36-45, 46-55, 56-65, and > 65. We first removed cases that had missing data for the question “What is the approximate age of the perpetrator?” in order to run the one-way ANOVA, Kruskal-Wallis, and Chi-Square Test of Independence analyses. **Table 1** shows the descriptive statistics and analyses for this reduced dataset (n=1,412).

**Table 1.**
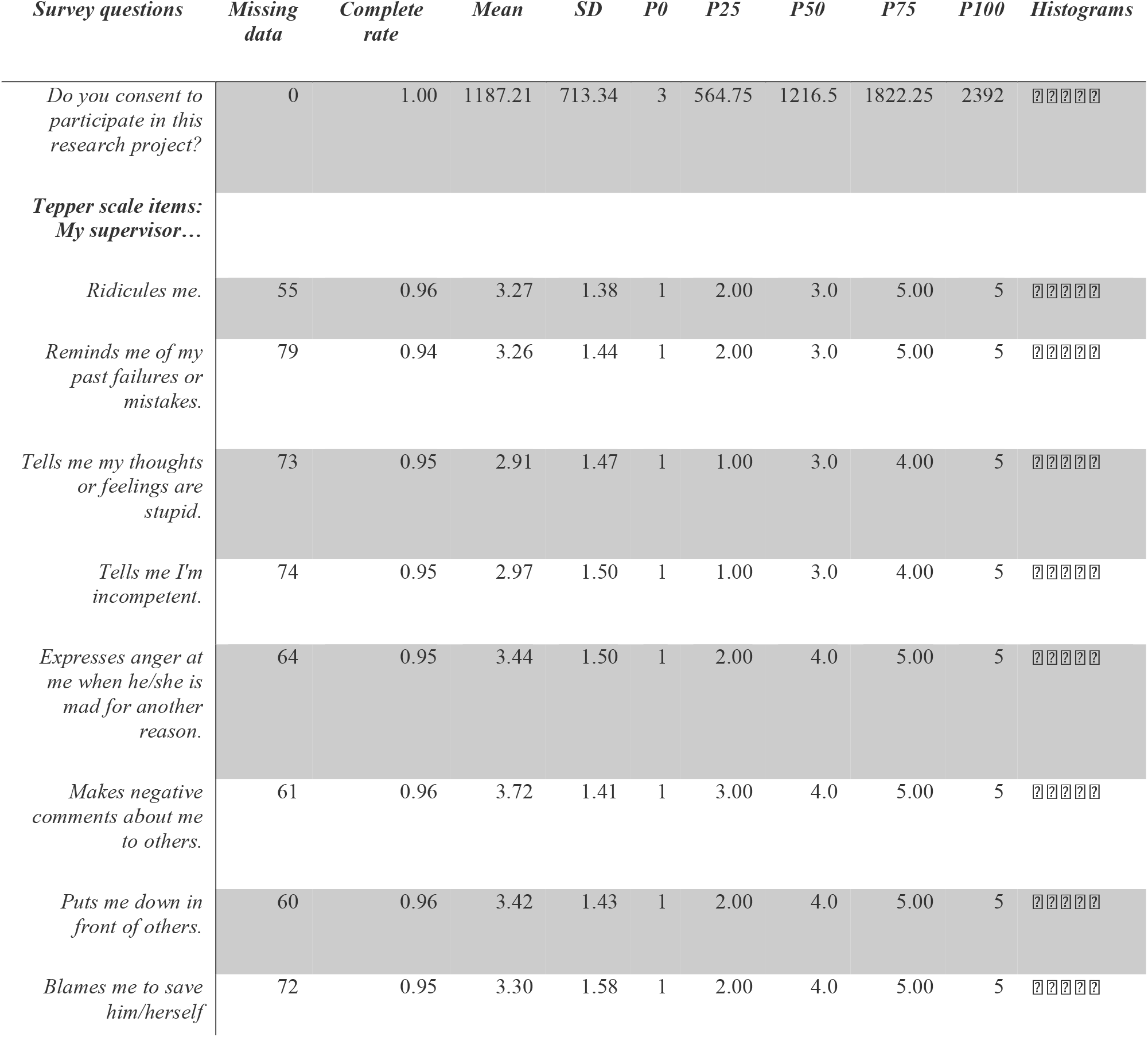
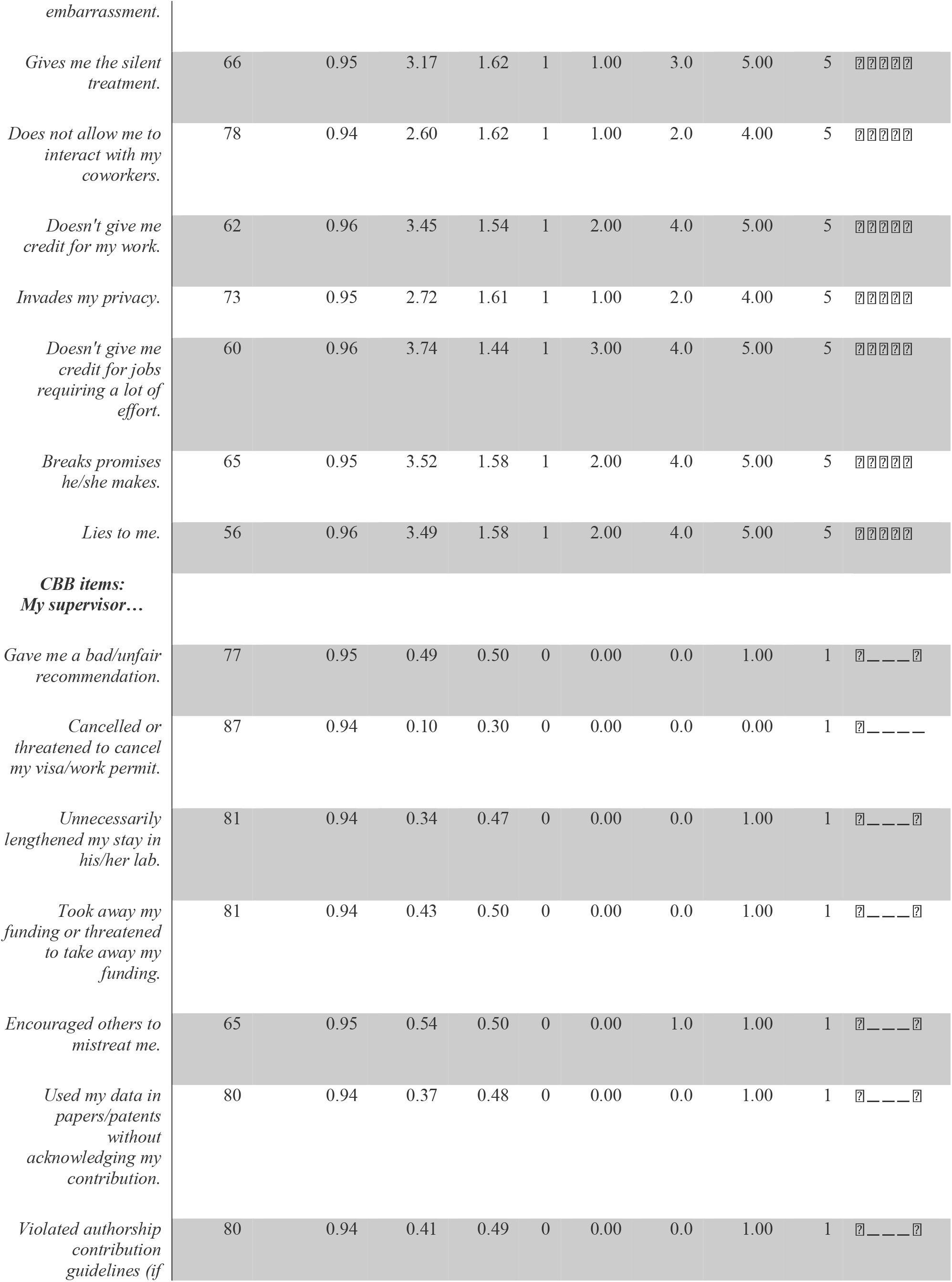
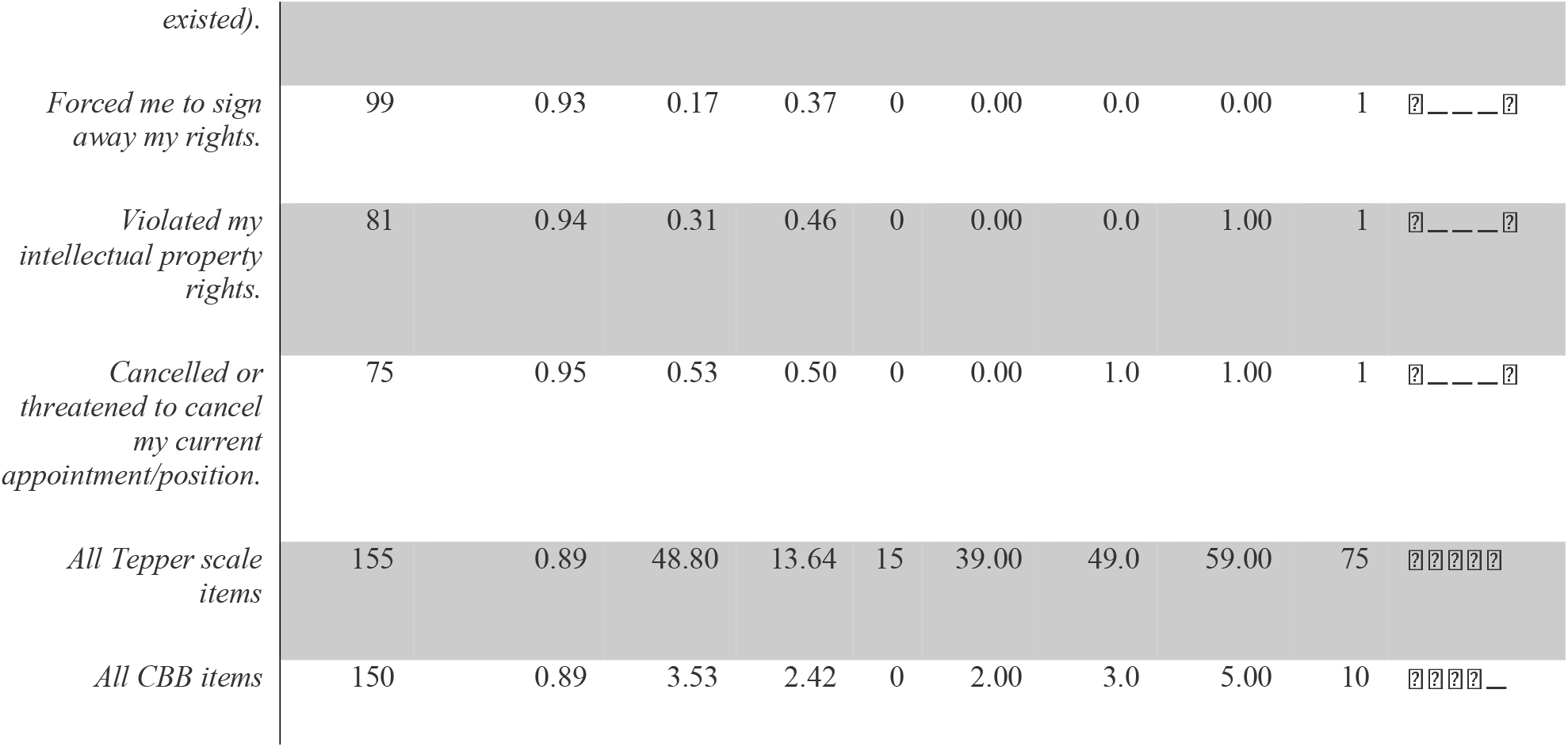
An overview of the dataset for the variables of interest. Age of perpetrator and academic bullying behaviors, defined as items on the Tepper Scale and Checklist of Bullying Behaviors (CBB). The survey questions details are available in **the Supporting Information**. (“P” stands for “Percentile”)

We used the 15-item abusive supervision scale developed by Tepper^18^ to assess general bullying behaviors. Sample items included statements like “my supervisor ridicules me” and “my supervisor puts me down in front of others.” Participants who perceived themselves as having been bullied responded to these items using a 5-point Likert scale ranging from 1 (“I cannot remember him/her ever using this behavior with me”) to 5 (“He/she uses this behavior very often with me”). In addition to the Tepper scale, we utilized our contextual bullying behavior (CBB) checklist, which consists of 10 items, and was developed through the analysis of publicized stories and narratives collected over time by the authors.^5^ These behaviors include abusing authorship or violating intellectual property rights^19^; threatening to cancel funding, positions, or visas^6^; and damaging the reputations of budding scientists through negative recommendations or speaking poorly about them to others^12^. It is noteworthy that we linked the study of what is colloquially known as “bullying” in academic science with the established science of abusive supervision from the organizational literature.^5^

To determine whether the age of perpetrators affects their level of abusive supervision, we investigated the relationship between age categories from the survey and scores on the Tepper^18^ scale, which measures abusive supervision, together with the influence of age on the contextual behaviors associated with abusive actions. We ran analyses looking at the entire Tepper scale (15 items). The Tepper scale score distribution across the age groups was similar enough to use a one-way ANOVA analysis to compare the total Tepper scale score by perpetrator age. 155 responses were removed from the analysis due to missing data. We did not find a significant difference among any of the perpetrator age groups and total Tepper scale score (**Figure 1a-d**). Full data analysis of each individual behavior with perpetrator age group is available on the **SI**.

**Figure 1.**
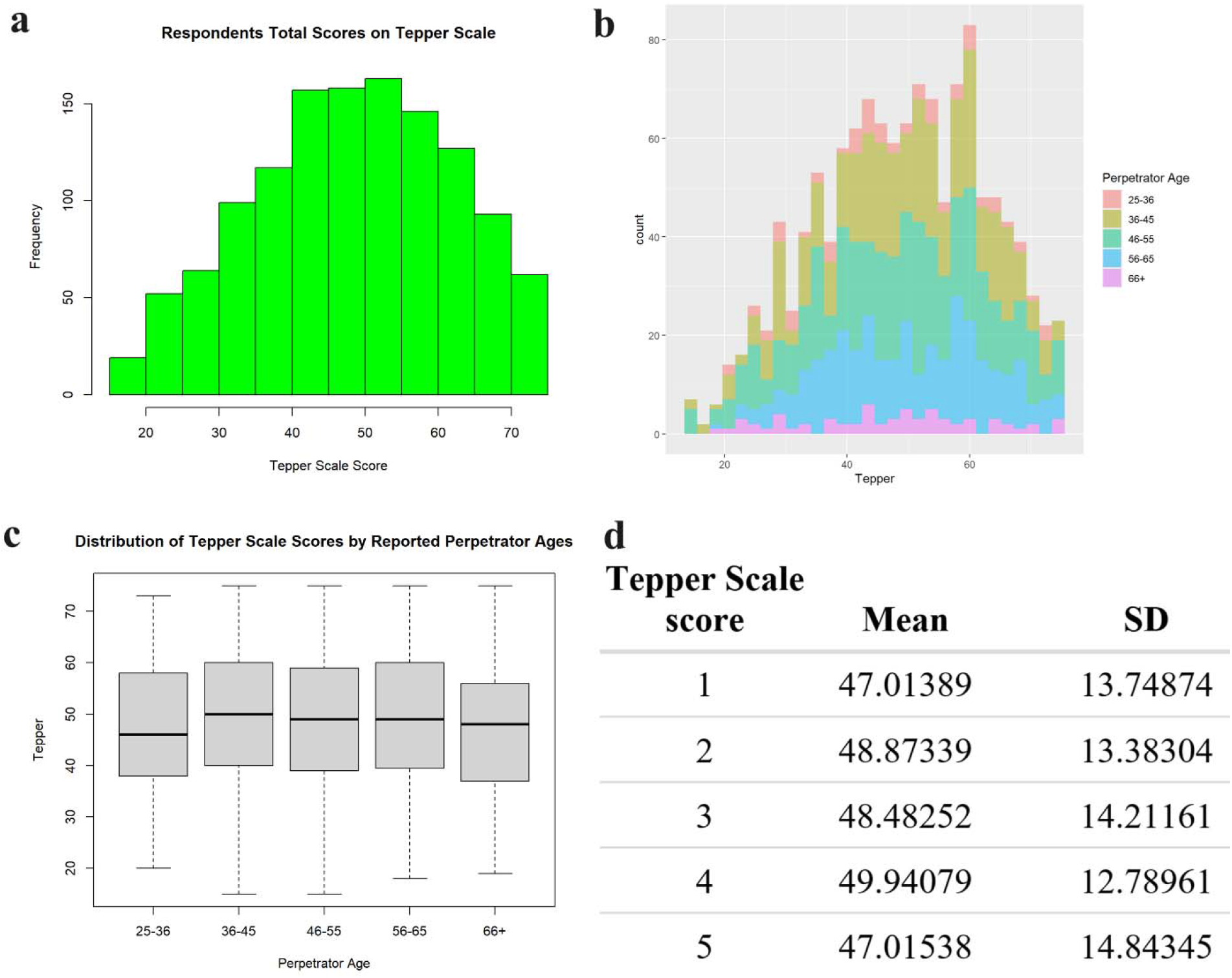
Tepper scale analysis of the age of the perpetrators. Frequency (a) and counts (b) of the respondents of the total scores on the Tepper scale; (c) box plots showing the distribution of the Tepper scale scores by the reported perpetrator ages; (d) one-way ANOVA results comparing perpetrator age group to Tepper Scale score.

**Figure 2.**
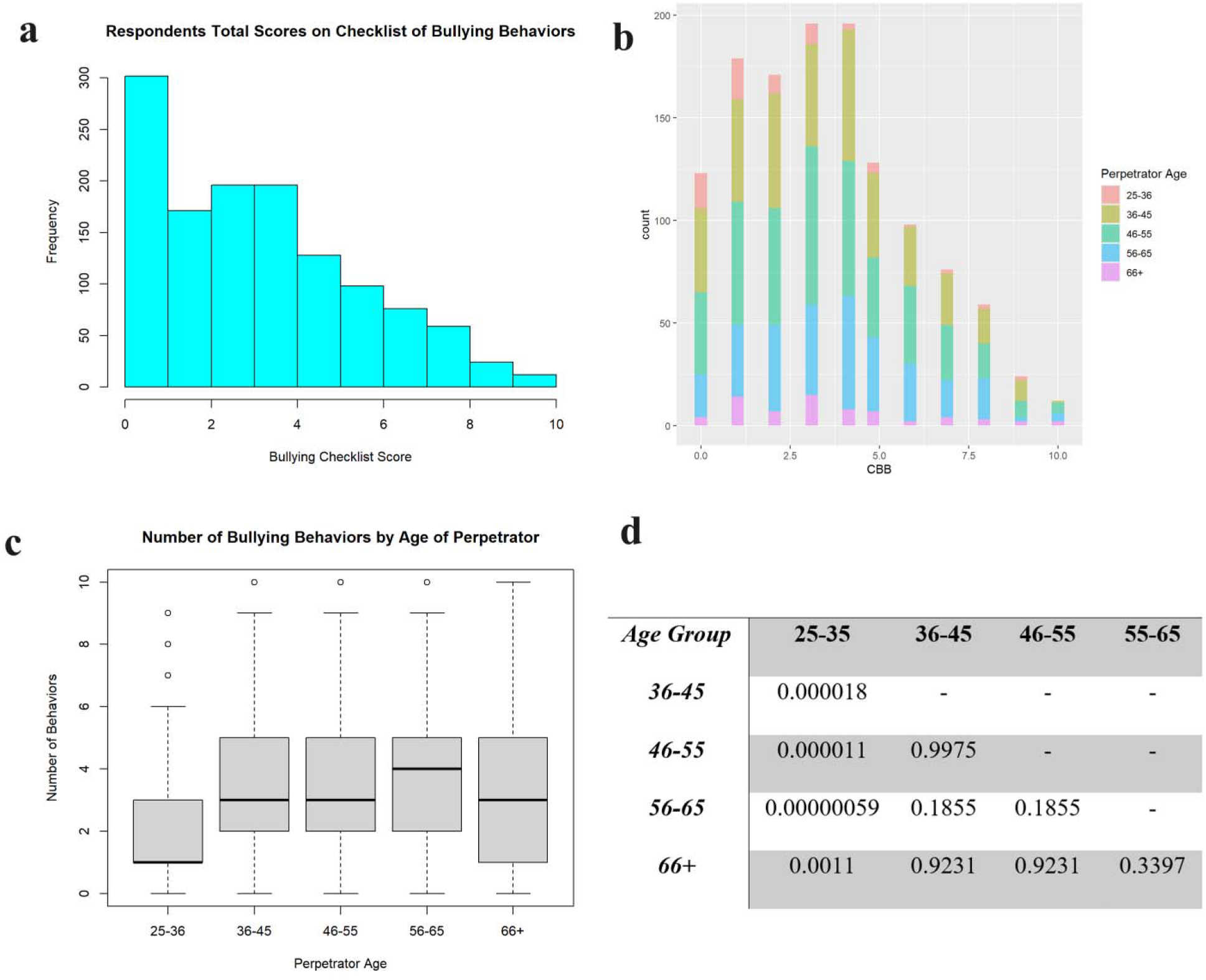
Contextual bullying behavior (CBB) checklist analysis of the age of the perpetrators. Frequency (a) and counts (b) of the respondents of the total scores on the CBB; (c) box plots showing the distribution of the number of abusive behaviors by the reported perpetrator ages; and (d) pairwise comparisons using Wilcoxon rank sun test with continuity correction. Age group “25-35” is significantly different than all other age groups.

We then investigated whether the perpetrator’s academic position, ethnicity, and sex influenced the observed age-related results. Our analysis revealed no significant interactions between the perpetrator’s academic position, ethnicity, or sex with their age group. Specifically, the professional positions of the perpetrators—Principal Investigator (n=722), Lab Supervisor (n=75), Group Leader (n=155), and Department Chair (n=239)—showed no significant correlation with the perpetrator’s age groups. Furthermore, there was no additive effect between the perpetrator’s age group and their professional role, as indicated by the non-significant results from the unbalanced two-way ANOVA.

Similarly, no significant interactions were found between the perpetrator’s age group and their race/ethnicity—White (n=860), Asian (n=133), Middle Eastern (n=65), and East Indian (n=40). However, an additive relationship between the perpetrator’s age group and their race/ethnicity was observed, with the unbalanced two-way ANOVA indicating a significant result (p<0.01). Tukey’s HSD test identified one significant relationship between the “White” and “Asian” groups. Nonetheless, post-hoc analyses revealed that the ANOVA normality assumption was not met (p<0.001), as the Shapiro-Wilk normality test result was below the threshold of p<0.05.

Additionally, there were no interactions between the perpetrator’s age group and sex. An additive relationship was found between the perpetrator’s age group and sex—male (n=751) and female (n=420)—with the unbalanced two-way ANOVA showing a significant result (p<0.01). However, post-hoc analyses revealed that ANOVA normality assumption was not met (p<0.001). To be met, the Shapiro-Wilk normality test must not be p<0.05.

Overall, these analyses suggest that the perpetrator’s age group results were not significantly influenced by their academic position, ethnicity, or sex.

Next, we ran analyses looking at the summed checklist of binary yes/no academic bullying behaviors (10 items) as well as the individual items in relation to the reported age of the perpetrator (**Table 2**). The distribution of the CBB scores across the age groups were not similar enough to use a one-way ANOVA analysis. Because these were skewed, we used a Kruskal-Wallis test to compare the total CBB scale score by perpetrator age. 150 responses were removed from the analysis due to missing data. We found a significant difference among the lowest perpetrator age groups (25-35) and total CBB score when compared to all other perpetrator age groups (p<0.01).

**Table 2.**
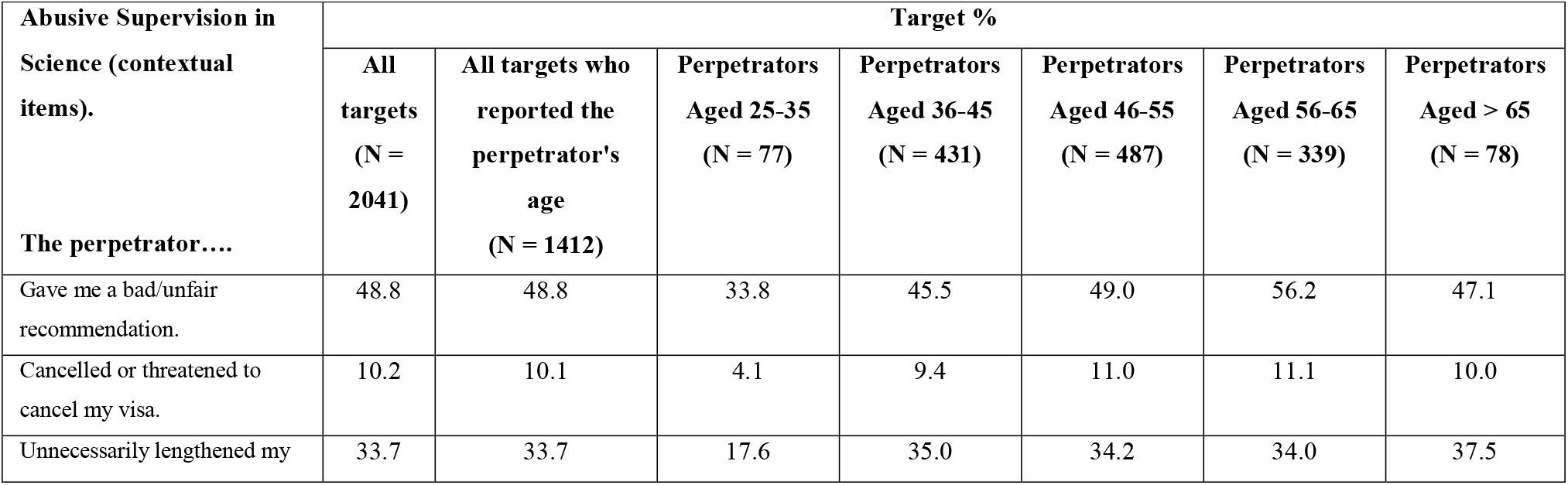
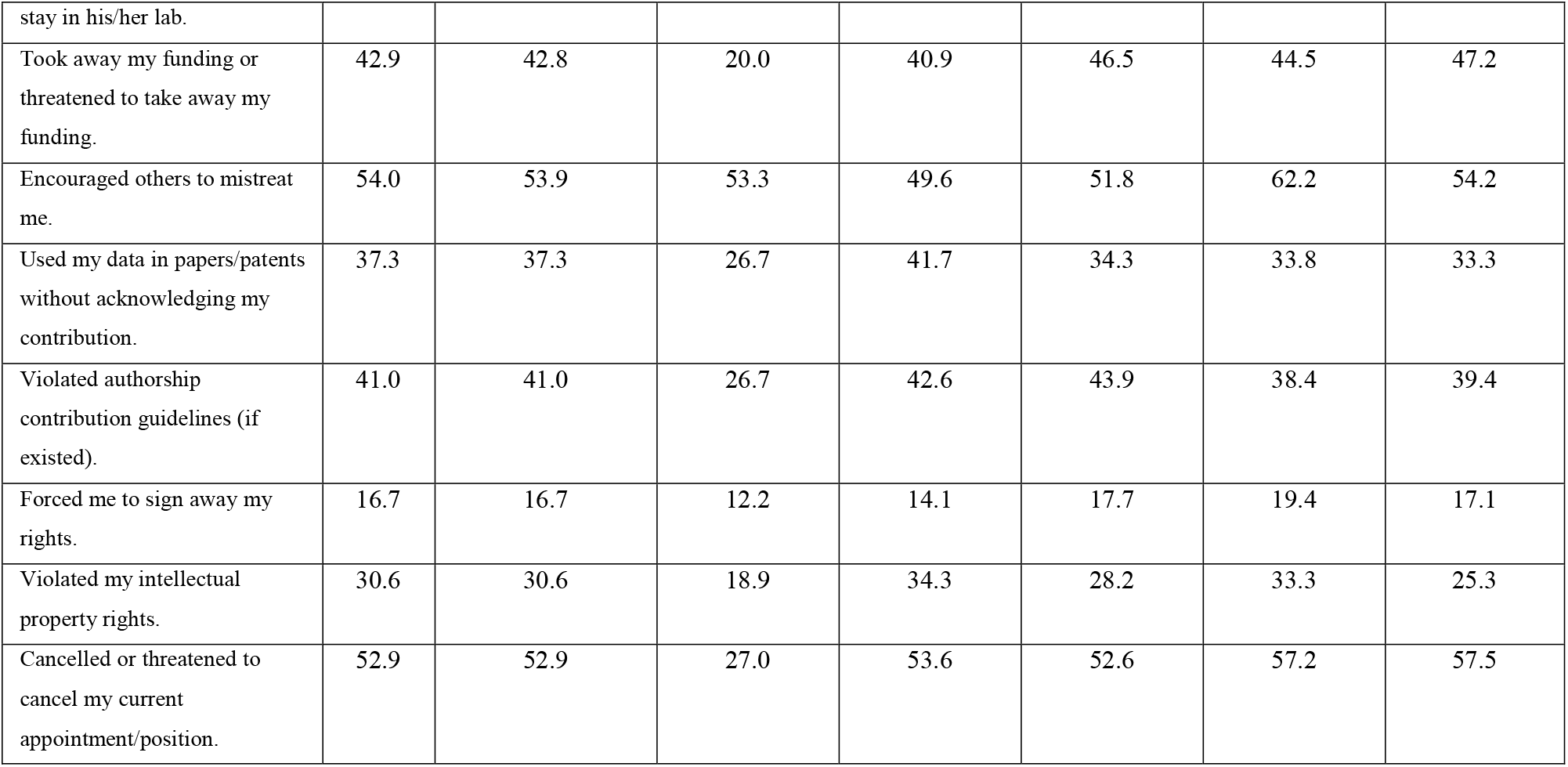
Percentage of perpetrators engaging in contextual bullying behaviors across age groups.

We finally looked at the individual items on the CBB in relation to the perpetrator’s age group. We ran chi-square tests of independence to analyze these items, as both the predictor variable (perpetrator age) and outcome variable (CBB item) were categorical variables (**Table 3** and **Figure 3;** full details of the statistical analysis are available in the **SI**). We found five items with a significant relationship between the bullying behavior and perpetrator age group (p<0.05) and 3 items with less significant differences (0.05<p<0.1). The two items lacking a significant relationship were: “Cancelled or threatened to cancel by visa/work permit” and “Forced me to sign away my rights.” Both of these CBBs appear to be low-frequency behaviors relative to the other CBBs, possibly because they are more extreme. It’s also interesting that younger perpetrators were as likely as most other age groups to encourage others to mistreat targets. While this is egregious behavior, it is less likely observable, traceable, or punishable compared to the more serious behaviors (e.g., “violate IP or violating authorship rights”). Interestingly, the age group that stands out as more likely to write unfair recommendation letters and encourage others to mistreat targets is 56-65.

**Table 3.** Contextual bullying behaviors with a significant overall difference between age groups. See additional data for the trends of all 10 contextual bullying behaviors across age-groups.

| Contextual Bullying Behavior | Chi-Square | df | p-value | Specific Differences** |
| --- | --- | --- | --- | --- |
| Gave me a bad/unfair recommendation | 15.510 | 4 | $p < 0.004$ | 25-35 (lower), $p < .01$<br>56-65 (higher), $p < .002$ |
| Unnecessarily lengthened my stay in lab | 9.427 | 4 | $p < 0.051^*$ | 25-35 (lower) |
| Took away funding or threatened to do so | 19.125 | 4 | $p < 0.001$ | 25-35 (lower), $p < .0001$ |
| Encouraged others to mistreat me | 13.213 | 4 | $p < 0.01$ | 56-65 (higher), $p < .0005$ |
| Used my data/patents | 9.170 | 4 | $p < 0.057^*$ | 36-45 (higher), $p < .02^*$ |
| Violated authorship contribution guidelines | 8.844 | 4 | $p < 0.065^*$ | 25-35 (lower), $p < .009$ |
| Violated my intellectual property rights | 11.201 | 4 | $p < 0.024$ | 25-35 (lower), $p < .028^*$ |
| Cancelled/Threatened to cancel position | 23.006 | 4 | $p < 0.001$ | 25-35 (lower), $p < .0001$ |

\*Indicates significance slightly above the conventional .05 level; \*\* Based on post-hoc Chi-square tests with Bonferonni corrected p-values ( $p < 0.01$ is significant)

**Figure 3.**
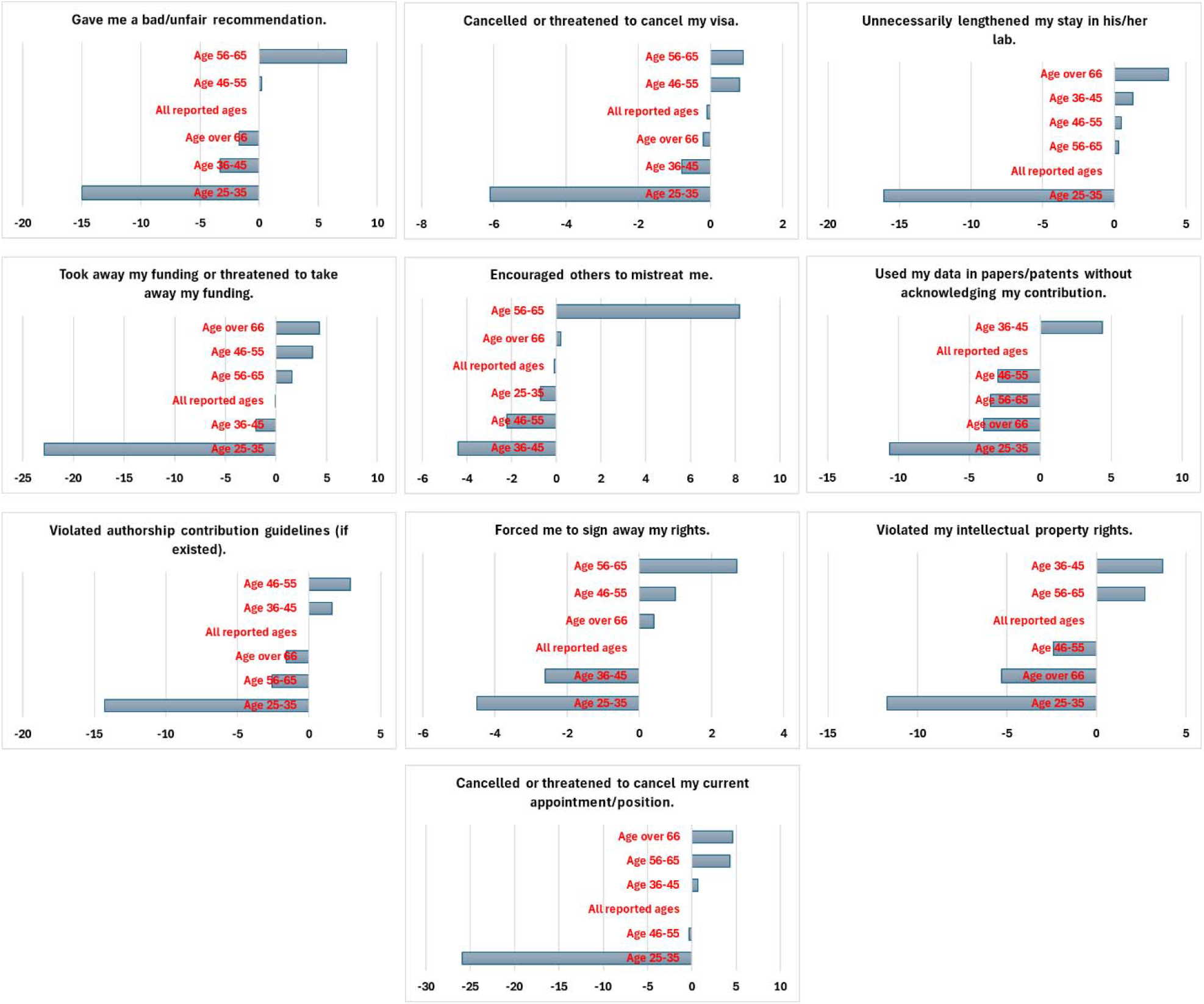
Effect of perpetrators’ age on the ten contextual abusive behaviors. Bar charts illustrating changes in perpetrators’ ten contextual abusive behaviors by age groups, with all percentages deducted from the “All Targets” category for comparison.

On the other hand, it appears that younger perpetrators are significantly less likely than the older groups to (1) give an unfair recommendation, (2) take away funding or threaten to, (3) violate targets’ authorship rights, and (4) threaten to cancel targets’ positions. Some of these CBBs (e.g. take away funding, violate authorship rights, cancelling targets’ positions/appointments) are serious behaviors a young, freshly minted principal investigators could be disciplined (e.g. denied tenure) or fired for, which may explain why they use these tactics to a lesser degree.

The findings from this study highlight the significant mental health benefits of addressing and mitigating academic bullying, particularly in relation to the age of perpetrators. Academic bullying has long been recognized as a detrimental factor that negatively impacts the mental well-being of targets and the people in the circle of their influence.^4,20-23^ By identifying that younger perpetrators (aged 25-35) are less likely to engage in abusive behaviors compared to their older counterparts, this research opens the door to more targeted interventions that can prevent the escalation of bullying behaviors over time. Early intervention, specifically aimed at younger faculty members or supervisors, could play a crucial role in reducing the prevalence of bullying and fostering a more supportive academic environment.

The study’s implications extend beyond merely identifying patterns of bullying behavior across different age groups. By understanding that older perpetrators are more likely to engage in a broader range of abusive behaviors, academic institutions can develop age-specific training programs that address these tendencies. This proactive approach could significantly alleviate the mental health burdens faced by victims of bullying, as it would work to reduce the frequency and severity of such incidents. Moreover, by curbing the behaviors of younger perpetrators early on, institutions can potentially prevent the long-term psychological damage that often results from prolonged exposure to academic bullying.

In addition to the direct mental health benefits for targets, the study also highlights the importance of creating a healthier and more inclusive academic environment, which can have broader positive effects on the entire academic community.^4^ When bullying behaviors are effectively managed and minimized, it not only benefits the immediate targets but also fosters a culture of respect and collaboration. This, in turn, can enhance overall job satisfaction, reduce stress levels, and contribute to the mental well-being of all members of the academic community. Furthermore, a more supportive environment can encourage greater innovation and productivity, as individuals feel more valued and less threatened, leading to a more vibrant and successful academic institution.

In summary, our cross-sectional global survey of academic bullying revealed that the age of perpetrators significantly influences their contextual abusive behaviors. Our results showed that the youngest perpetrators (aged 25-35) were less abusive compared to other age groups, exhibiting the lowest percentage of CBBs. Our data suggests that as perpetrators age, they will engage in a greater variety of CBBs, adding more CBBs to their repertoire of abuse as time goes by. This is strictly counter to the general trend which suggests that as people age, they become more prosocial and empathetic. These findings have strong implications for the development of more customized academic bullying training, policy development, and monitoring strategies. The most obvious one is to address the bullying behaviors of younger perpetrators early and severely to discourage them from continuing the behaviors and adding to their arsenals. Addressing age-specific dynamics in abusive behaviors can lead to more effective interventions and a healthier academic environment, ultimately benefiting the scientific community and its progress.

## Data Availability

All data produced in the present study are available upon reasonable request to the authors

