## Supplementary material for "Examining Age-Dependent Patterns in Academic Bullying Behaviors": SI

**Survey questions used in this analysis**

Q1 Do you consent to participate in this research project?

- Yes (1)
- No (0)

Q4 You have indicated that you have directly experienced academic bullying. Which of the following types of individuals was the perpetrator? If more than one, please choose one and refer to this person when answering the remaining questions in this section.

- Principal Investigator (1)
- Lab Supervisor (2)
- Group Leader (3)
- Department Chair (4)
- Senior Colleague (6)
- Head of a Public Research Institute or Observatory (9)
- Head of Private company research group or lab (10)
- Other (7)

Q5 If you answered "other" above, please fill in the type of perpetrator below.

________________________________________________________________

Q6 Please describe the perpetrator below.


Male or Female?

- Male (1)
- Female (2)
- Other (3)
- Prefer not to specify (4)

Q7 Was this person...

- White (1)
- Black or African American (2)
- American Indian or Alaska Native (3)
- Asian (4)
- Native Hawaiian or Pacific Islander (5)
- Hispanic (6)
- Mixed Race (7)
- Middle Eastern (8)
- East Indian (9)
- Other (10)

Q12 Please indicate the extent to which this person engages (or engaged if in the past) in the following behaviors toward you.

|  | I cannot remember him/her ever using this behavior with me (1) | He/she very seldom uses this behavior with me (2) | He/she occasionally uses this behavior with me (3) | He/she uses this behavior moderately often with me (4) | He/she uses this behavior very often with me (5) |
| --- | --- | --- | --- | --- | --- |
| Ridicules me. (1) |  |  |  |  |  |
| Reminds me of my past failures or mistakes. (2) |  |  |  |  |  |
| Tells me my thoughts or feelings are stupid. (3) |  |  |  |  |  |
| Tells me I'm incompetent. (4) |  |  |  |  |  |
| Expresses anger at me when he/she is mad for another reason. (5) |  |  |  |  |  |
| Makes negative comments about me to others. (6) |  |  |  |  |  |
| Puts me down in front of others. (7) |  |  |  |  |  |
| Blames me to save him/herself embarrassment. (8) |  |  |  |  |  |
| Gives me the silent treatment. (9) |  |  |  |  |  |
| Does not allow me to interact with my coworkers. (10) |  |  |  |  |  |
| Doesn't give me credit for my work. (11) |  |  |  |  |  |
| Invades my privacy. (12) |  |  |  |  |  |
| Doesn't give me credit for jobs requiring a lot of effort (13) |  |  |  |  |  |
| Breaks promises he/she makes (14) |  |  |  |  |  |
| Lies to me (15) |  |  |  |  |  |

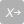

Q13 Has this person ever directed any of the following behaviors toward you?

|  | Yes (1) | No (0) |
| --- | --- | --- |
| Gave me a bad/unfair recommendation (1) |  |  |
| Cancelled or threatened to cancel my visa/work permit (2) |  |  |
| Unnecessarily lengthened my stay in his/her lab (3) |  |  |
| Took away my funding or threatened to take away my funding (4) |  |  |
| Encouraged others to mistreat me (5) |  |  |
| Used my data in papers/patents without acknowledging my contribution (6) |  |  |
| Violated authorship contribution guidelines (if existed) (7) |  |  |
| Forced me to sign away my rights (8) |  |  |
| Violated my intellectual property rights (9) |  |  |
| Cancelled or threatened to cancel my current appointment/position (10) |  |  |

**Data analysis of each individual behavior with perpetrator age group**

**12_1. Ridiculing**

| Frequency of Ridiculing Respondent by Age of Perpetrator | | | | | |
| --- | --- | --- | --- | --- | --- |
|  | **Never** | **Seldom** | **Occasional** | **Moderately Often** | **Very Often** |
| 25-36 | 11 | 6 | 17 | 22 | 19 |
| 36-45 | 60 | 54 | 111 | 81 | 106 |
| 46-55 | 83 | 72 | 111 | 91 | 112 |
| 56-65 | 40 | 46 | 86 | 67 | 91 |
| 66+ | 16 | 8 | 18 | 9 | 20 |

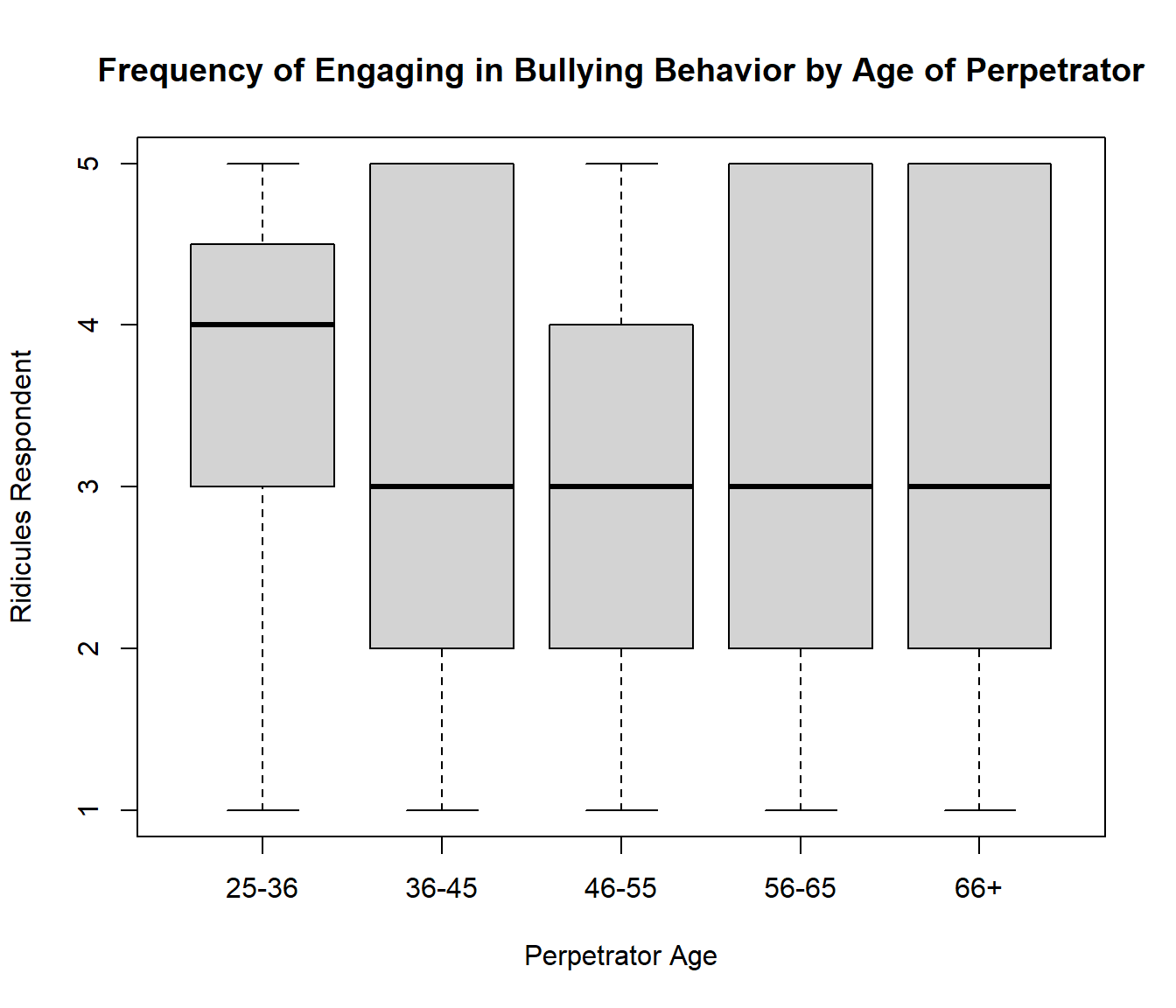

```
##
##  Pearson's Chi-squared test
##
## data:  table12_1
## X-squared = 17.33, df = 16, p-value = 0.3645
```

**12_2. Past Failures**

| Frequency of Reminding Respondent of Past Failures or Mistakes by Age of Perpetrator | | | | | |
| --- | --- | --- | --- | --- | --- |
|  | **Never** | **Seldom** | **Occasional** | **Moderately Often** | **Very Often** |
| 25-36 | 16 | 9 | 20 | 15 | 16 |
| 36-45 | 70 | 59 | 88 | 77 | 108 |
| 46-55 | 83 | 57 | 95 | 97 | 127 |
| 56-65 | 55 | 39 | 73 | 63 | 94 |
| 66+ | 15 | 10 | 12 | 15 | 20 |

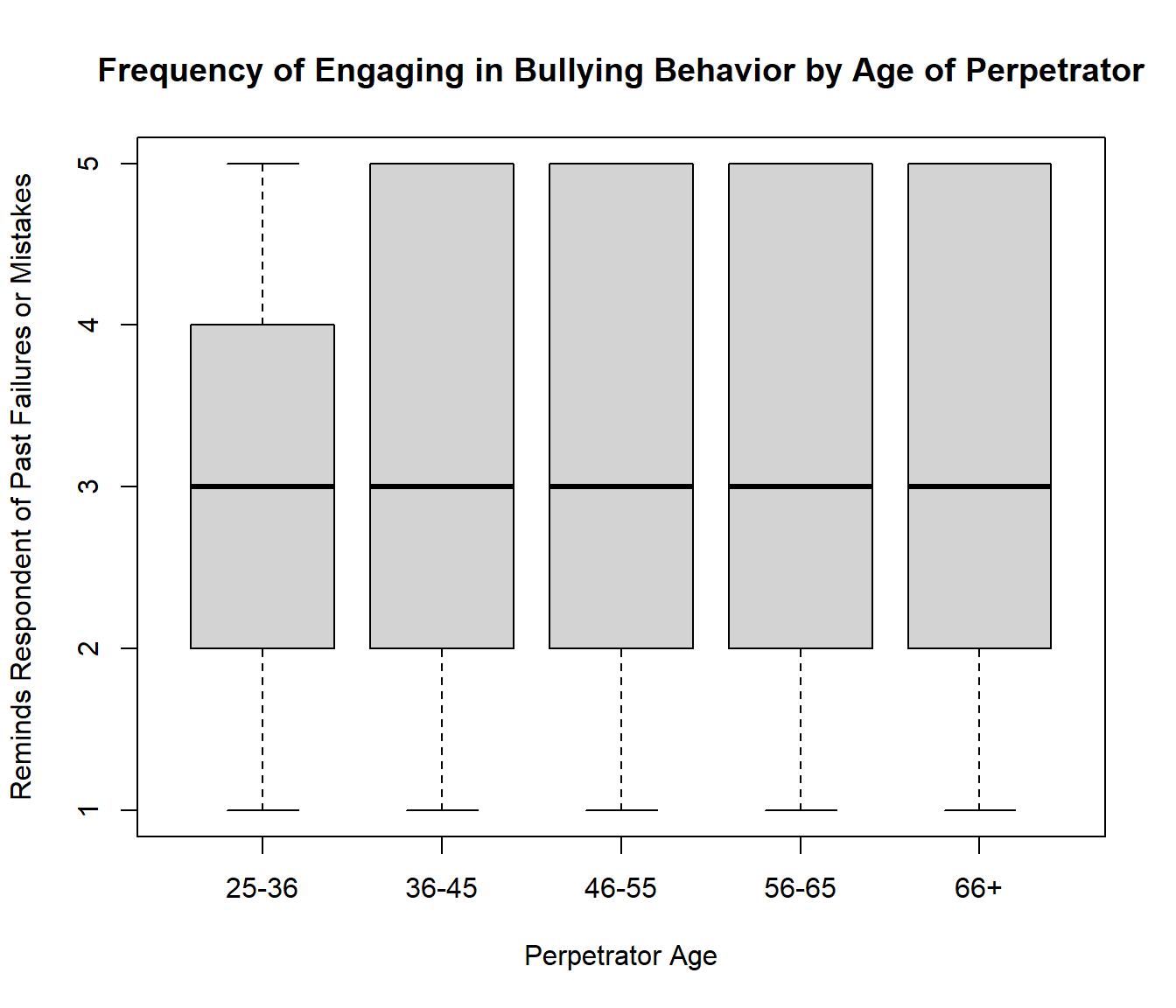

```
##
##  Pearson's Chi-squared test
##
## data:  table12_2
## X-squared = 6.2263, df = 16, p-value = 0.9855
```

**12_3. Thoughts or Feelings are Stupid**

| Frequency of Telling Respondent Thoughts/Feelings are Stupid by Age of Perpetrator | | | | | |
| --- | --- | --- | --- | --- | --- |
|  | **Never** | **Seldom** | **Occasional** | **Moderately Often** | **Very Often** |
| 25-36 | 15 | 15 | 19 | 13 | 13 |
| 36-45 | 110 | 70 | 76 | 68 | 84 |
| 46-55 | 122 | 69 | 103 | 82 | 84 |
| 56-65 | 81 | 57 | 49 | 69 | 70 |
| 66+ | 15 | 10 | 16 | 12 | 17 |

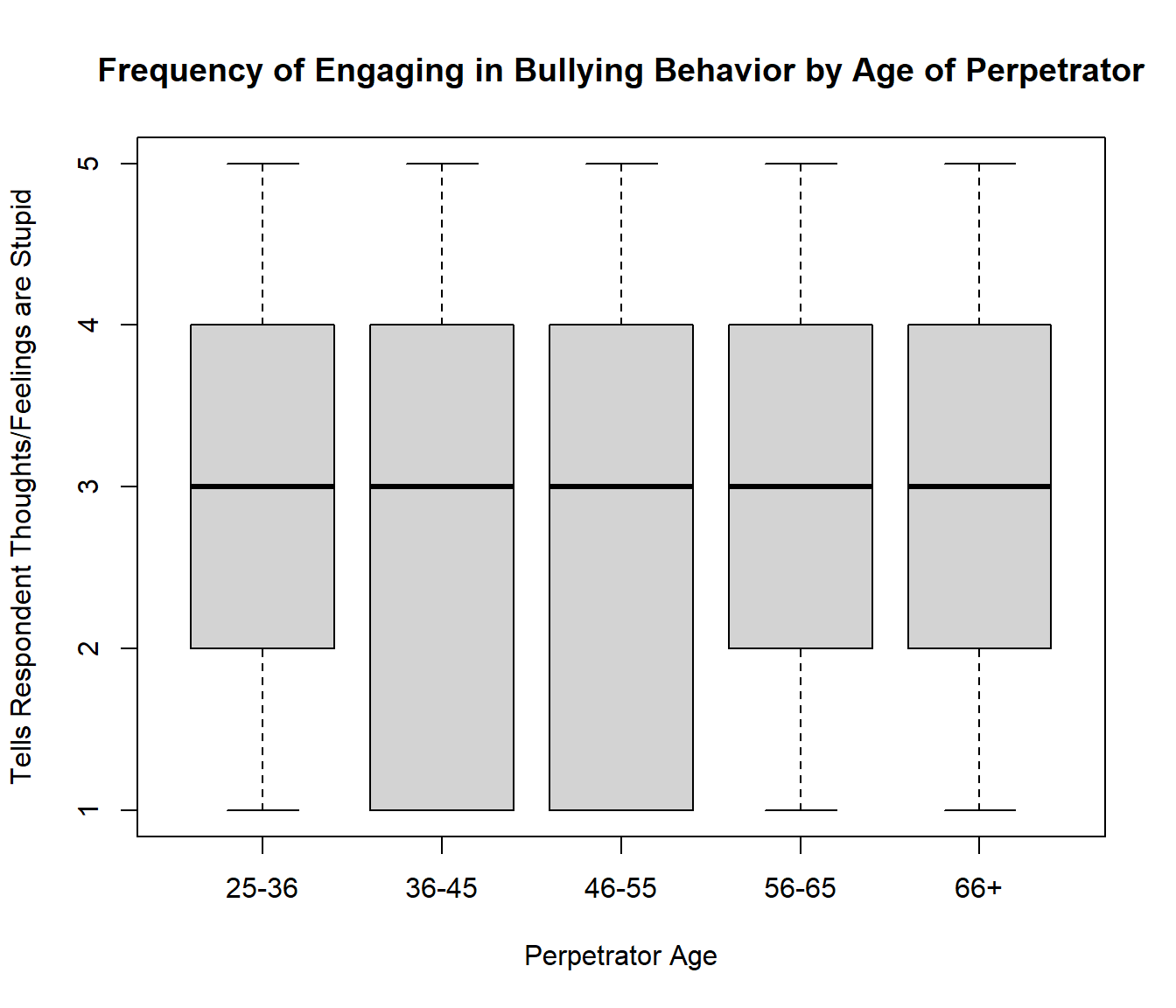

```
##
##  Pearson's Chi-squared test
##
## data:  table12_3
## X-squared = 14.958, df = 16, p-value = 0.5277
```

**12_4. Incompetent**

| Frequency of Telling Respondent They Are Incompetent by Age of Perpetrator | | | | | |
| --- | --- | --- | --- | --- | --- |
|  | **Never** | **Seldom** | **Occasional** | **Moderately Often** | **Very Often** |
| 25-36 | 18 | 12 | 11 | 18 | 17 |
| 36-45 | 100 | 72 | 71 | 71 | 92 |
| 46-55 | 124 | 75 | 80 | 76 | 101 |
| 56-65 | 81 | 36 | 70 | 67 | 74 |
| 66+ | 20 | 7 | 12 | 18 | 15 |

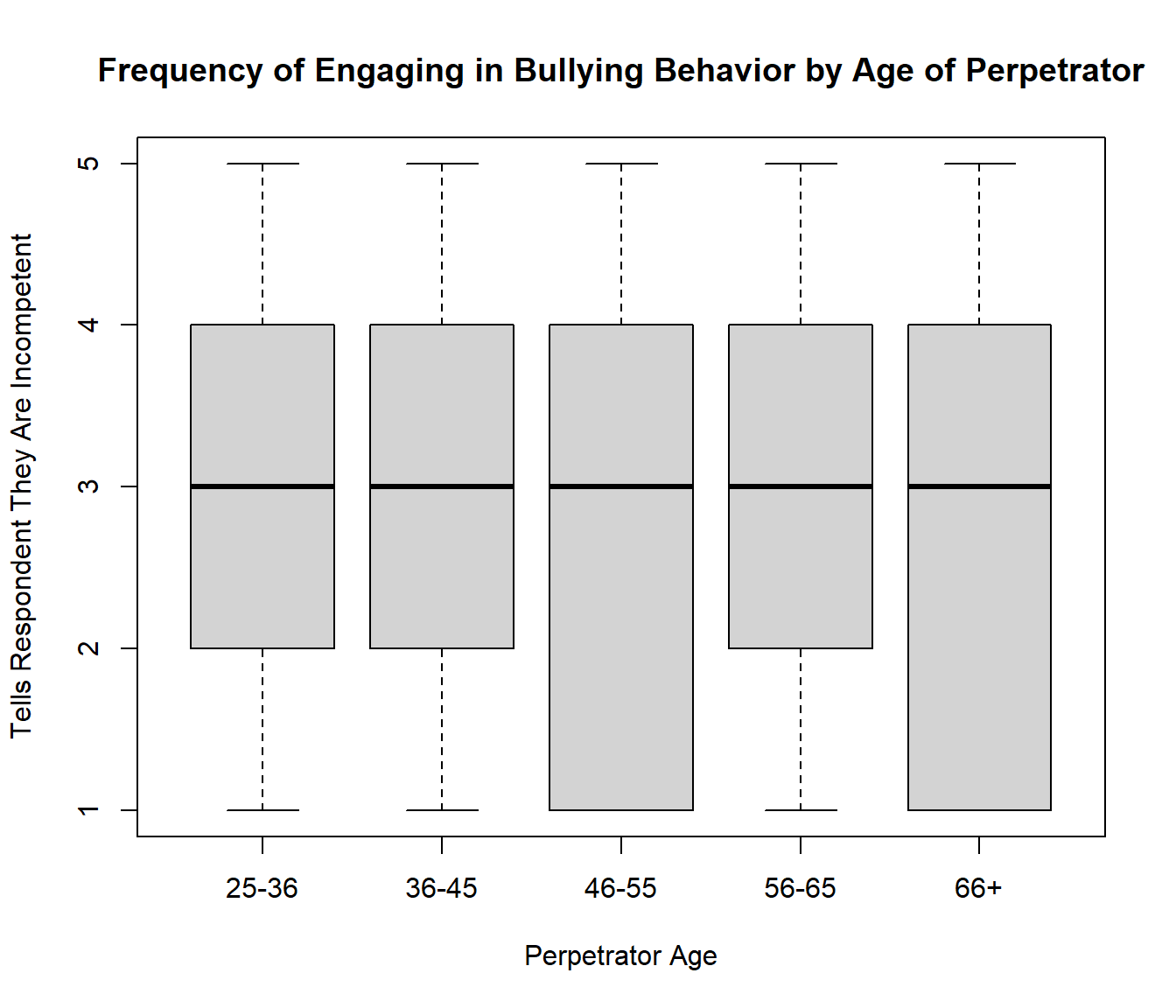

```
##
##  Pearson's Chi-squared test
##
## data:  table12_4
## X-squared = 15.623, df = 16, p-value = 0.4795
```

**12_5. Anger**

| Frequency of Expressing Anger Toward Respondent for Another Reason by Age of Perpetrator | | | | | |
| --- | --- | --- | --- | --- | --- |
|  | **Never** | **Seldom** | **Occasional** | **Moderately Often** | **Very Often** |
| 25-36 | 18 | 6 | 14 | 18 | 20 |
| 36-45 | 65 | 38 | 73 | 85 | 149 |
| 46-55 | 84 | 54 | 69 | 89 | 165 |
| 56-65 | 61 | 33 | 59 | 60 | 115 |
| 66+ | 16 | 5 | 10 | 13 | 29 |

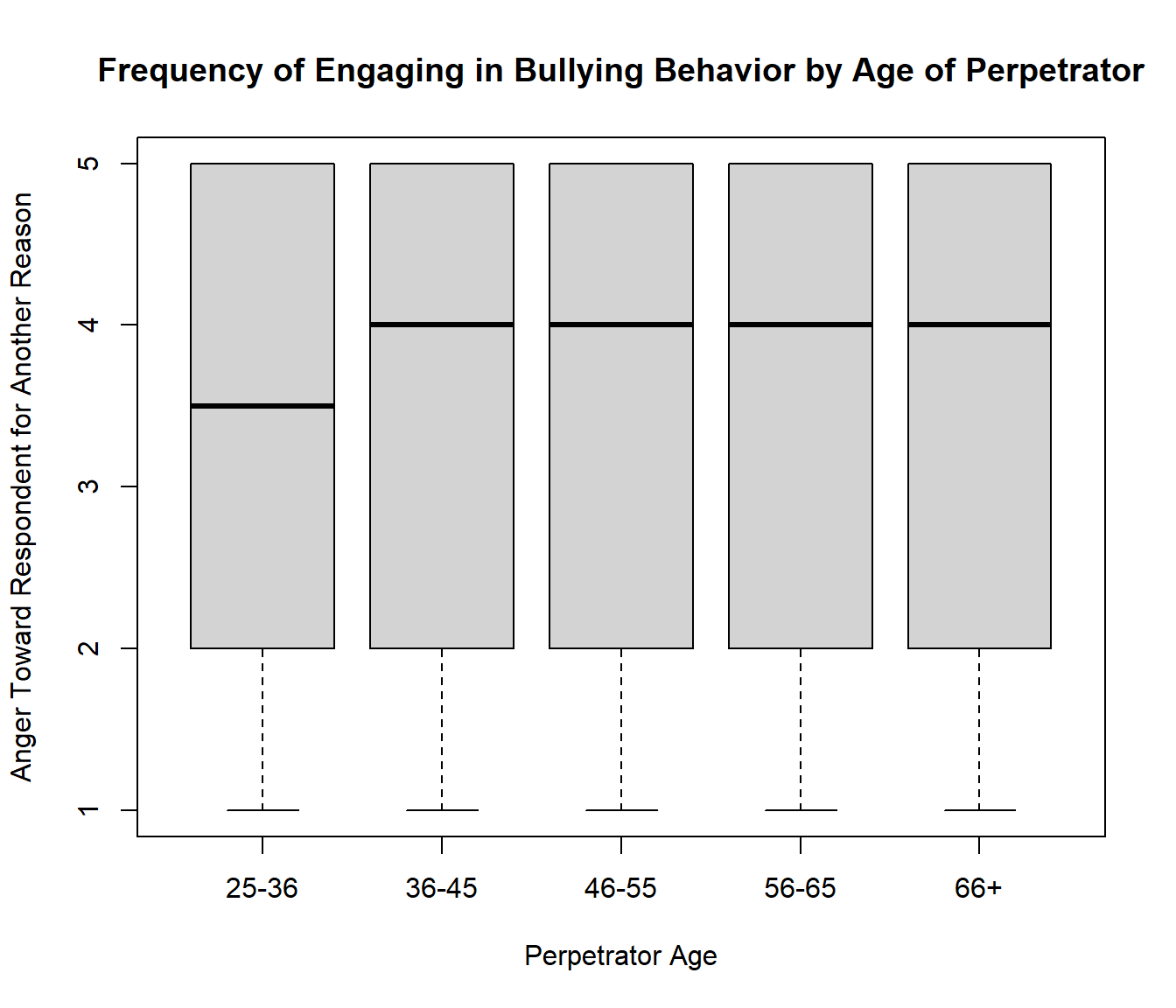

```
##
##  Pearson's Chi-squared test
##
## data:  table12_5
## X-squared = 11.294, df = 16, p-value = 0.791
```

**12_6. Negative Comments**

| Frequency of Making Negative Comments About Respondent to Others by Age of Perpetrator | | | | | |
| --- | --- | --- | --- | --- | --- |
|  | **Never** | **Seldom** | **Occasional** | **Moderately Often** | **Very Often** |
| 25-36 | 10 | 5 | 11 | 9 | 40 |
| 36-45 | 57 | 28 | 75 | 92 | 158 |
| 46-55 | 62 | 32 | 94 | 87 | 186 |
| 56-65 | 33 | 27 | 49 | 64 | 159 |
| 66+ | 13 | 6 | 8 | 15 | 31 |

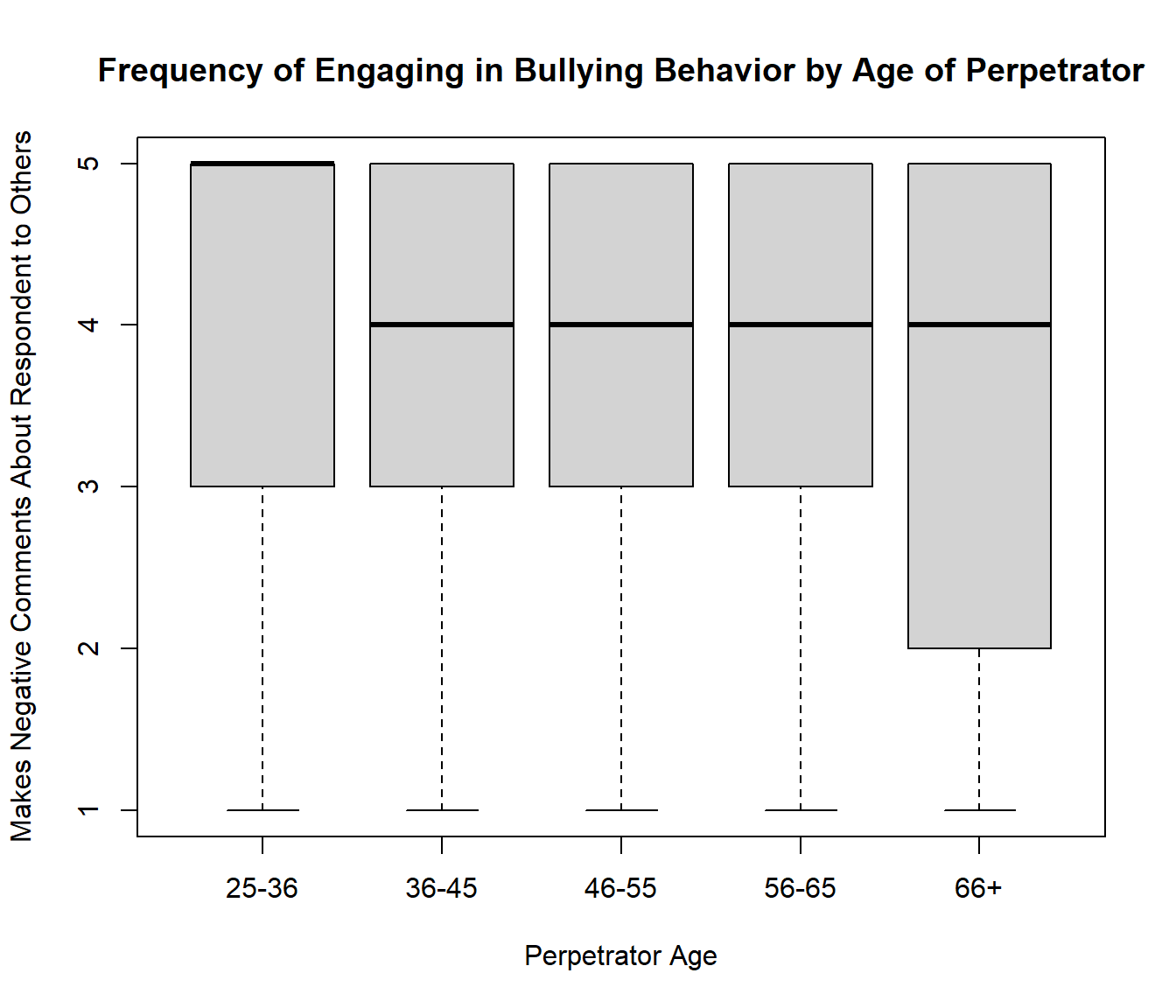

```
##
##  Pearson's Chi-squared test
##
## data:  table12_6
## X-squared = 20.952, df = 16, p-value = 0.1804
```

**12_7. Put Down**

| Frequency of Putting Down Respondent in Front of Others by Age of Perpetrator | | | | | |
| --- | --- | --- | --- | --- | --- |
|  | **Never** | **Seldom** | **Occasional** | **Moderately Often** | **Very Often** |
| 25-36 | 10 | 14 | 14 | 16 | 22 |
| 36-45 | 57 | 65 | 85 | 86 | 119 |
| 46-55 | 66 | 65 | 96 | 82 | 151 |
| 56-65 | 53 | 32 | 48 | 74 | 124 |
| 66+ | 13 | 9 | 16 | 12 | 23 |

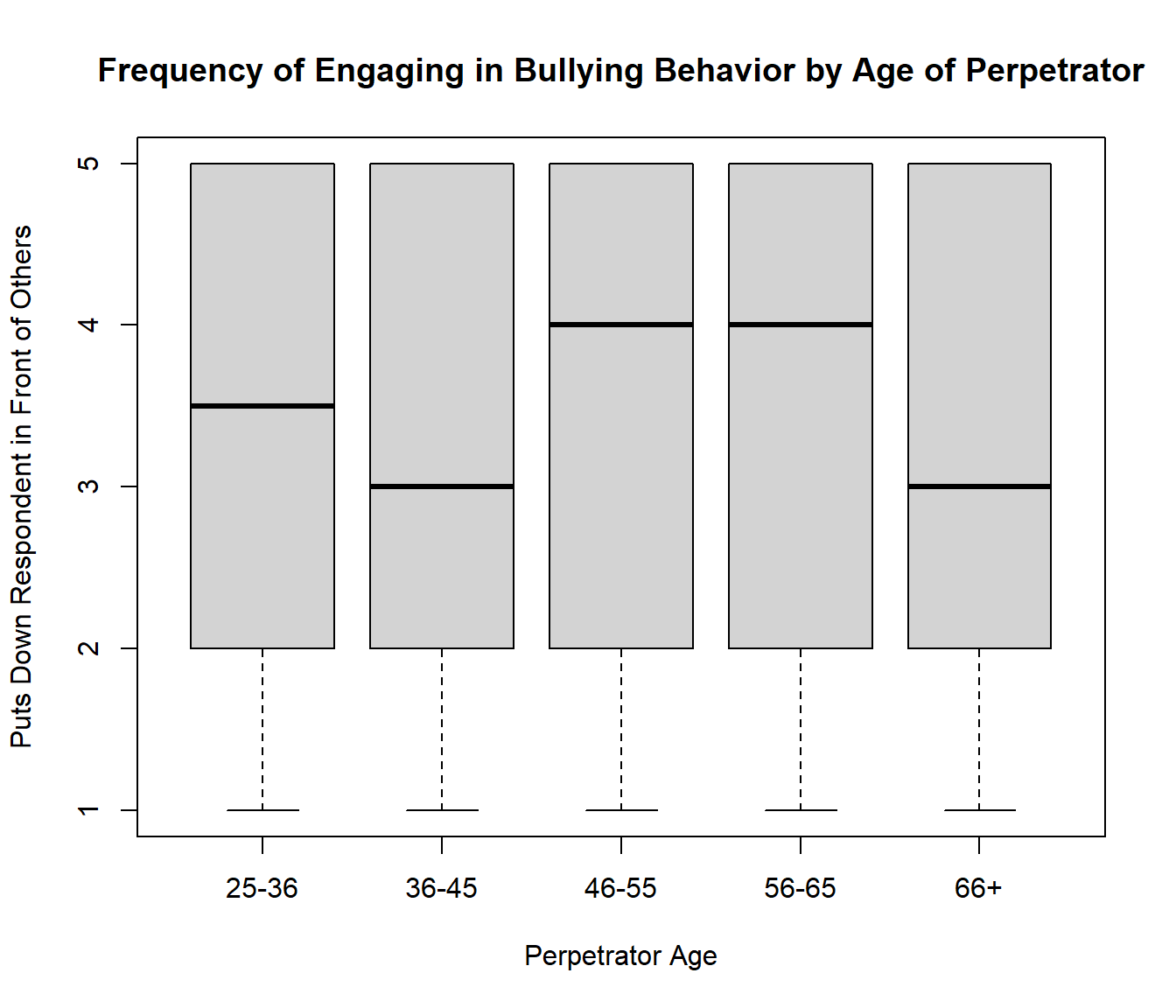

```
##
##  Pearson's Chi-squared test
##
## data:  table12_7
## X-squared = 20.271, df = 16, p-value = 0.2083
```

**12_8. Blame**

| Frequency of Blaming Respondent To Save Self Embarrassment by Age of Perpetrator | | | | | |
| --- | --- | --- | --- | --- | --- |
|  | **Never** | **Seldom** | **Occasional** | **Moderately Often** | **Very Often** |
| 25-36 | 26 | 8 | 7 | 14 | 21 |
| 36-45 | 84 | 44 | 70 | 70 | 141 |
| 46-55 | 105 | 51 | 63 | 77 | 163 |
| 56-65 | 72 | 30 | 44 | 66 | 114 |
| 66+ | 26 | 6 | 6 | 7 | 25 |

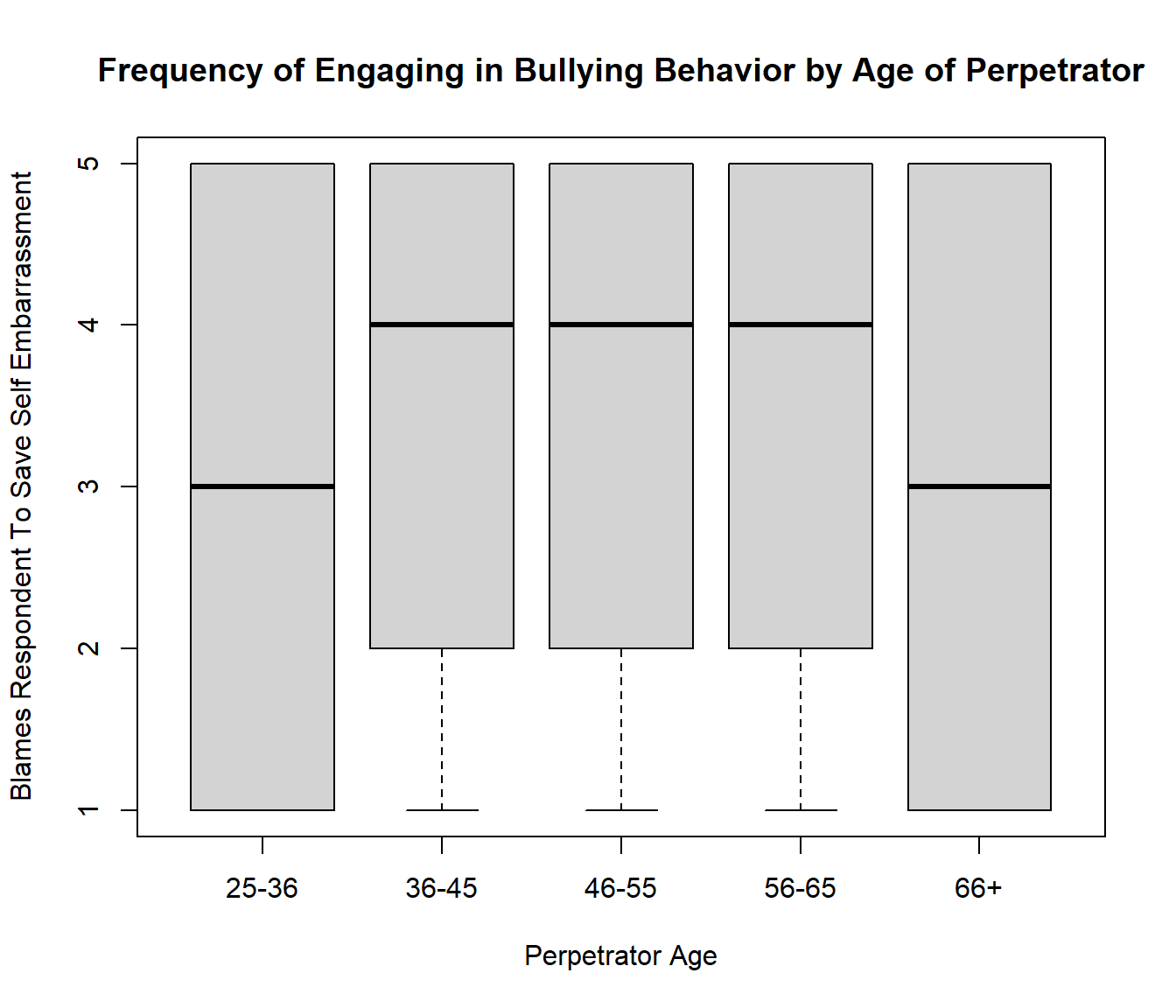

```
##
##  Pearson's Chi-squared test
##
## data:  table12_8
## X-squared = 22.763, df = 16, p-value = 0.1202
```

**12_9. Silent Treatment**

| Frequency of Giving Respondent Silent Treatment by Age of Perpetrator | | | | | |
| --- | --- | --- | --- | --- | --- |
|  | **Never** | **Seldom** | **Occasional** | **Moderately Often** | **Very Often** |
| 25-36 | 18 | 8 | 13 | 7 | 30 |
| 36-45 | 105 | 56 | 55 | 67 | 125 |
| 46-55 | 121 | 63 | 55 | 67 | 155 |
| 56-65 | 84 | 43 | 44 | 44 | 113 |
| 66+ | 20 | 5 | 9 | 12 | 27 |

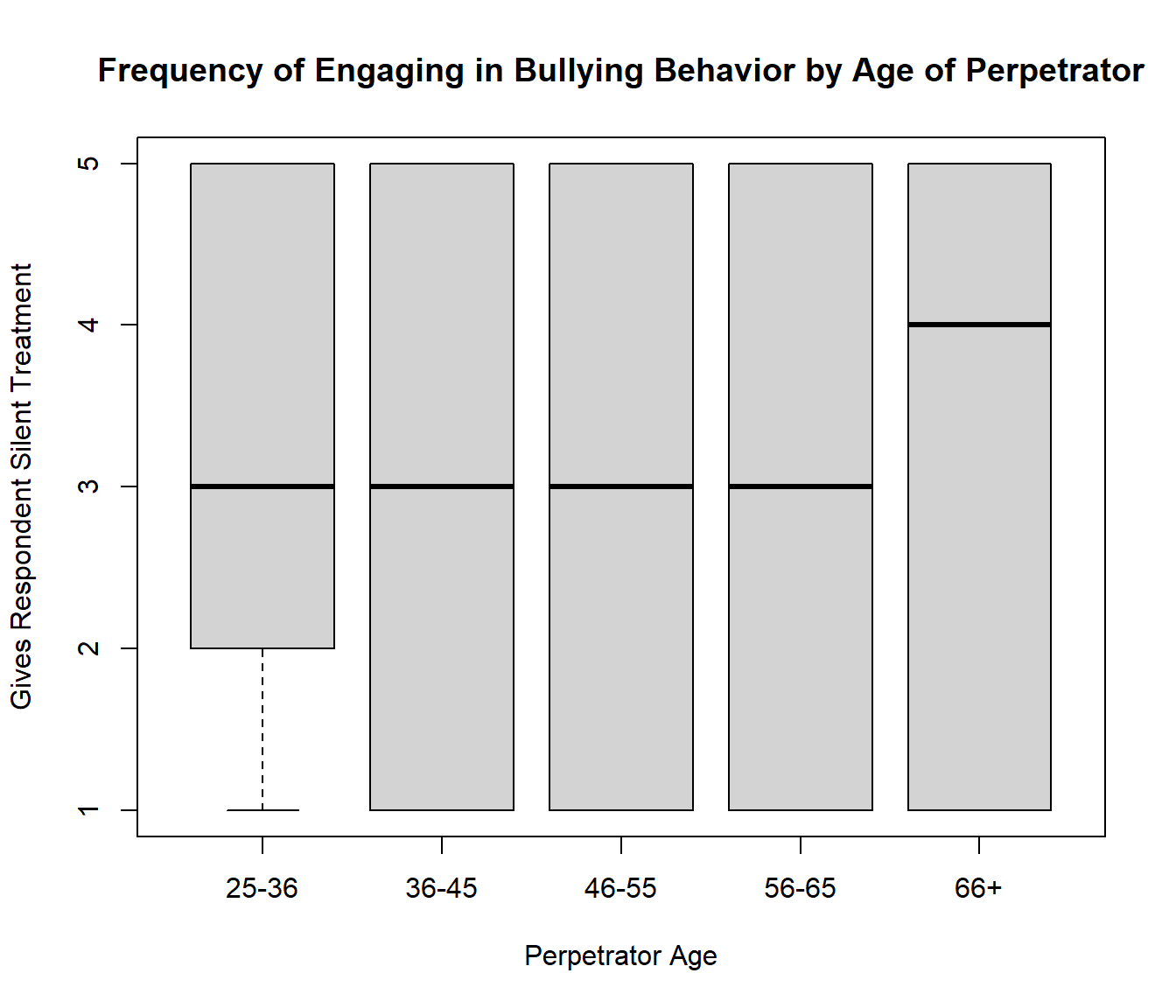

```
##
##  Pearson's Chi-squared test
##
## data:  table12_9
## X-squared = 9.65, df = 16, p-value = 0.8843
```

**12_10. No Interactions**

| Frequency of Not Letting Respondent Interact with Coworkers by Age of Perpetrator | | | | | |
| --- | --- | --- | --- | --- | --- |
|  | **Never** | **Seldom** | **Occasional** | **Moderately Often** | **Very Often** |
| 25-36 | 38 | 9 | 10 | 5 | 14 |
| 36-45 | 166 | 50 | 47 | 53 | 90 |
| 46-55 | 196 | 71 | 46 | 42 | 99 |
| 56-65 | 116 | 49 | 45 | 39 | 78 |
| 66+ | 32 | 8 | 5 | 6 | 20 |

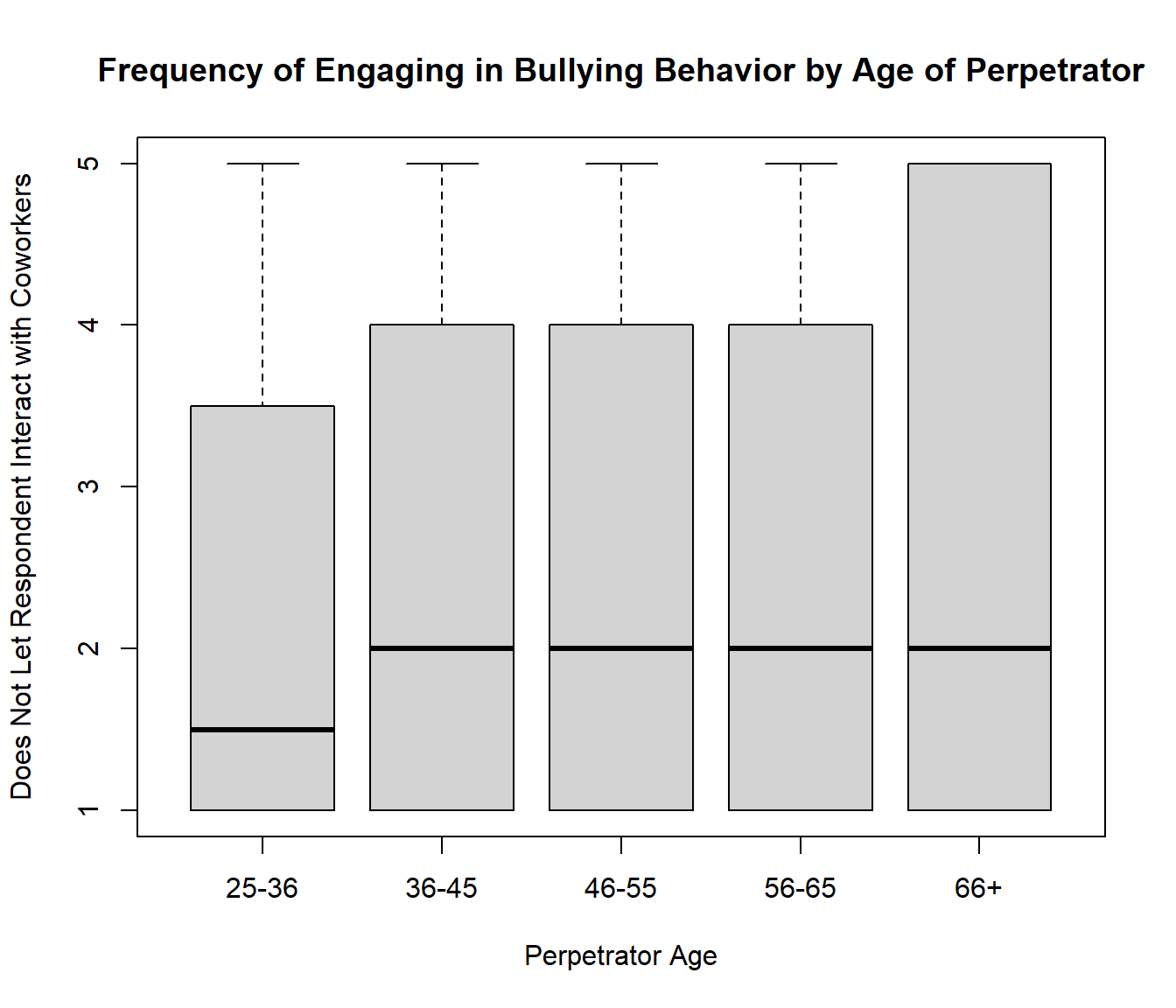

```
##
##  Pearson's Chi-squared test
##
## data:  table12_10
## X-squared = 17.73, df = 16, p-value = 0.3399
```

**12_11. No Credit for Work**

| Frequency of Not Giving Respondent Credit for Work by Age of Perpetrator | | | | | |
| --- | --- | --- | --- | --- | --- |
|  | **Never** | **Seldom** | **Occasional** | **ModeratelyOften** | **VeryOften** |
| 25-36 | 21 | 8 | 11 | 11 | 26 |
| 36-45 | 73 | 43 | 63 | 82 | 150 |
| 46-55 | 95 | 56 | 54 | 78 | 179 |
| 56-65 | 51 | 38 | 49 | 56 | 133 |
| 66+ | 19 | 4 | 12 | 9 | 29 |

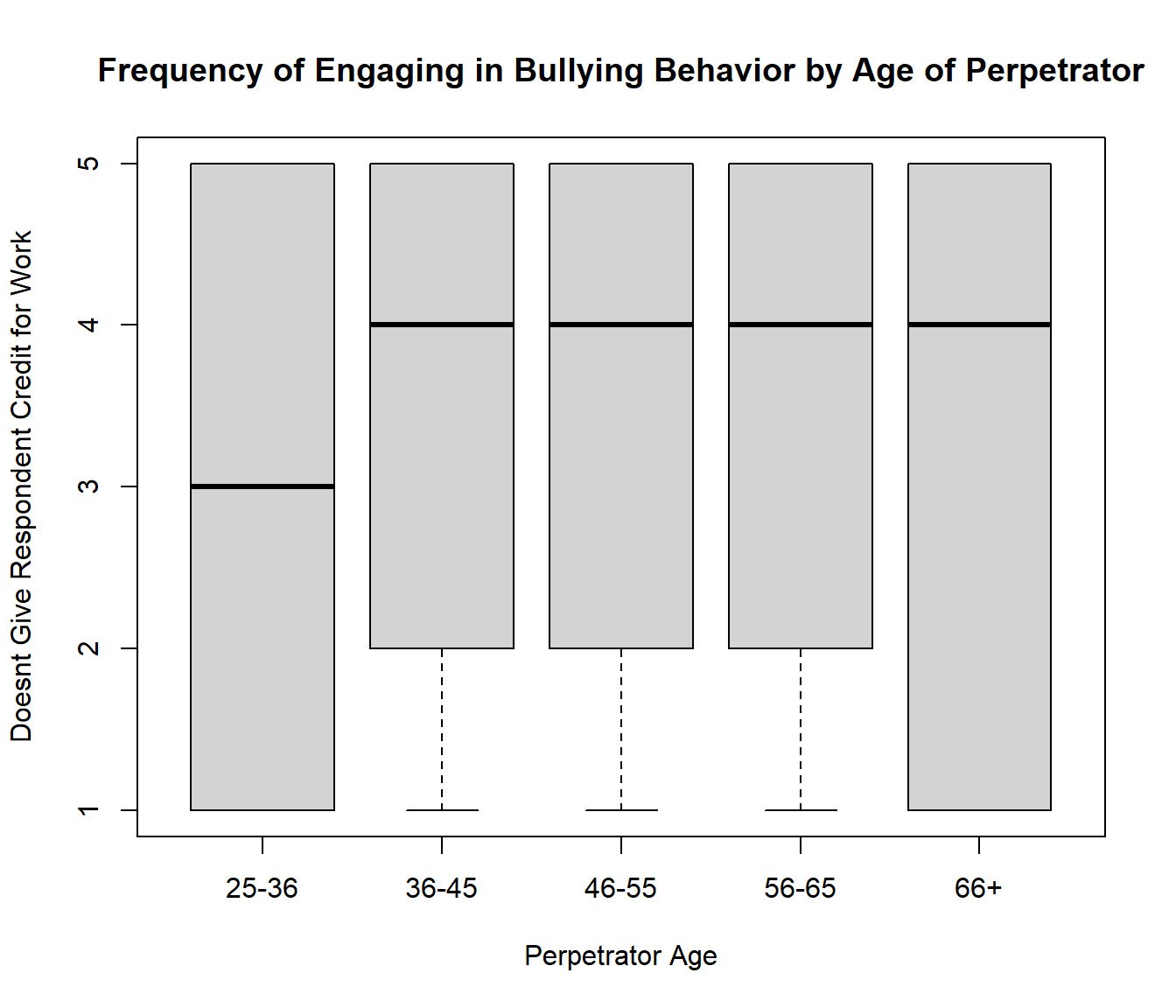

```
##
##  Pearson's Chi-squared test
##
## data:  table12_11
## X-squared = 17.521, df = 16, p-value = 0.3527
```

**12_12. Invade Privacy**

| Frequency of Invading Respondents Privacy by Age of Perpetrator | | | | | |
| --- | --- | --- | --- | --- | --- |
|  | **Never** | **Seldom** | **Occasional** | **Moderately Often** | **Very Often** |
| 25-36 | 27 | 10 | 6 | 11 | 22 |
| 36-45 | 135 | 52 | 73 | 58 | 90 |
| 46-55 | 172 | 64 | 47 | 69 | 105 |
| 56-65 | 124 | 47 | 47 | 33 | 75 |
| 66+ | 34 | 8 | 7 | 6 | 17 |

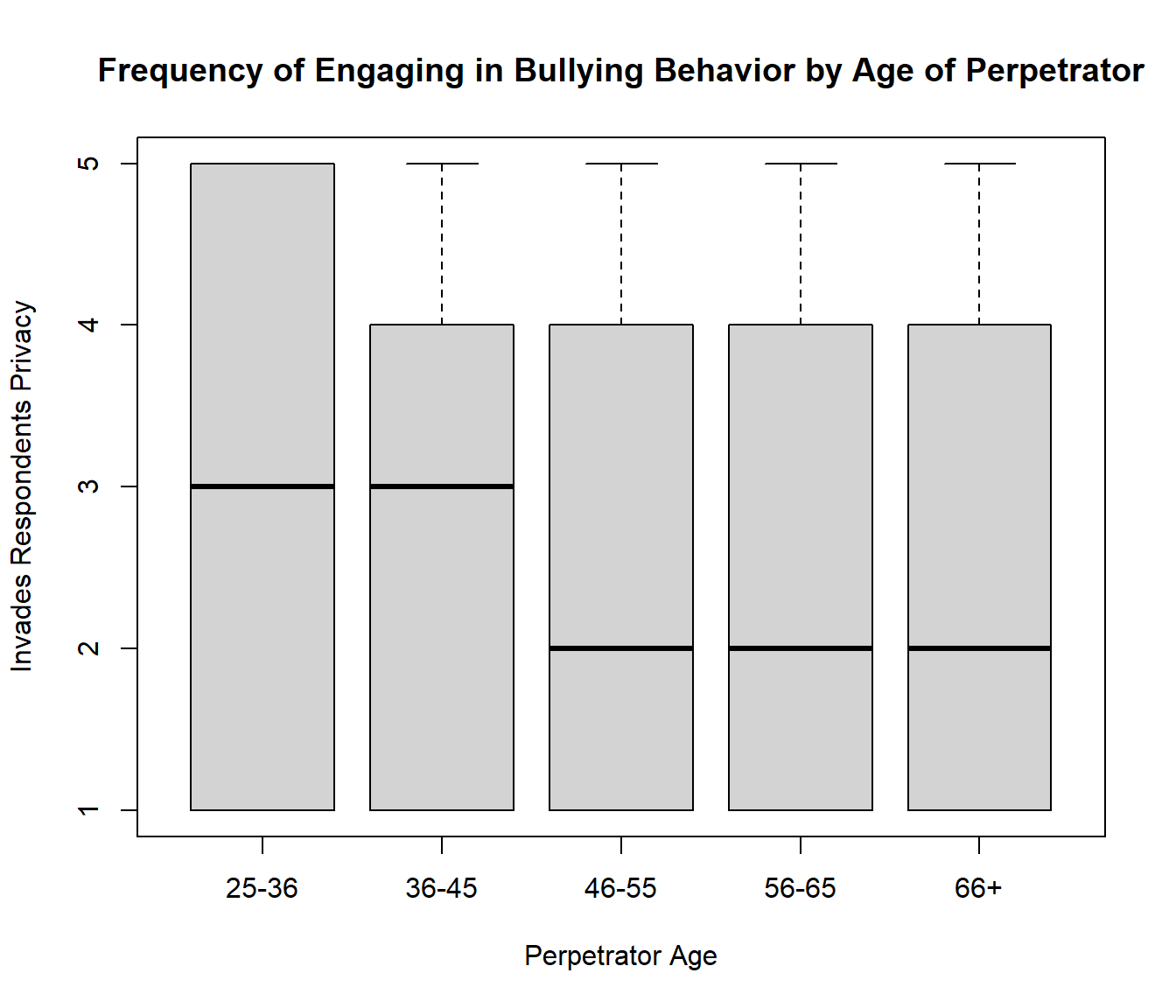

```
##
##  Pearson's Chi-squared test
##
## data:  table12_12
## X-squared = 23.409, df = 16, p-value = 0.1032
```

**12_13. No Credit for Effort**

There is a significant relationship (p=0.1) between the bullying behavior “not giving respondent credit for high effort jobs” and perpetrator age groups “25-35” and “56-65.”

| Frequency of Not Giving Respondent Credit for High Effort Jobs by Age of Perpetrator | | | | | |
| --- | --- | --- | --- | --- | --- |
|  | **Never** | **Seldom** | **Occasional** | **Moderately Often** | **Very Often** |
| 25-36 | 16 | 12 | 8 | 15 | 26 |
| 36-45 | 57 | 35 | 48 | 99 | 174 |
| 46-55 | 70 | 51 | 52 | 74 | 215 |
| 56-65 | 25 | 27 | 50 | 68 | 159 |
| 66+ | 8 | 7 | 15 | 6 | 35 |

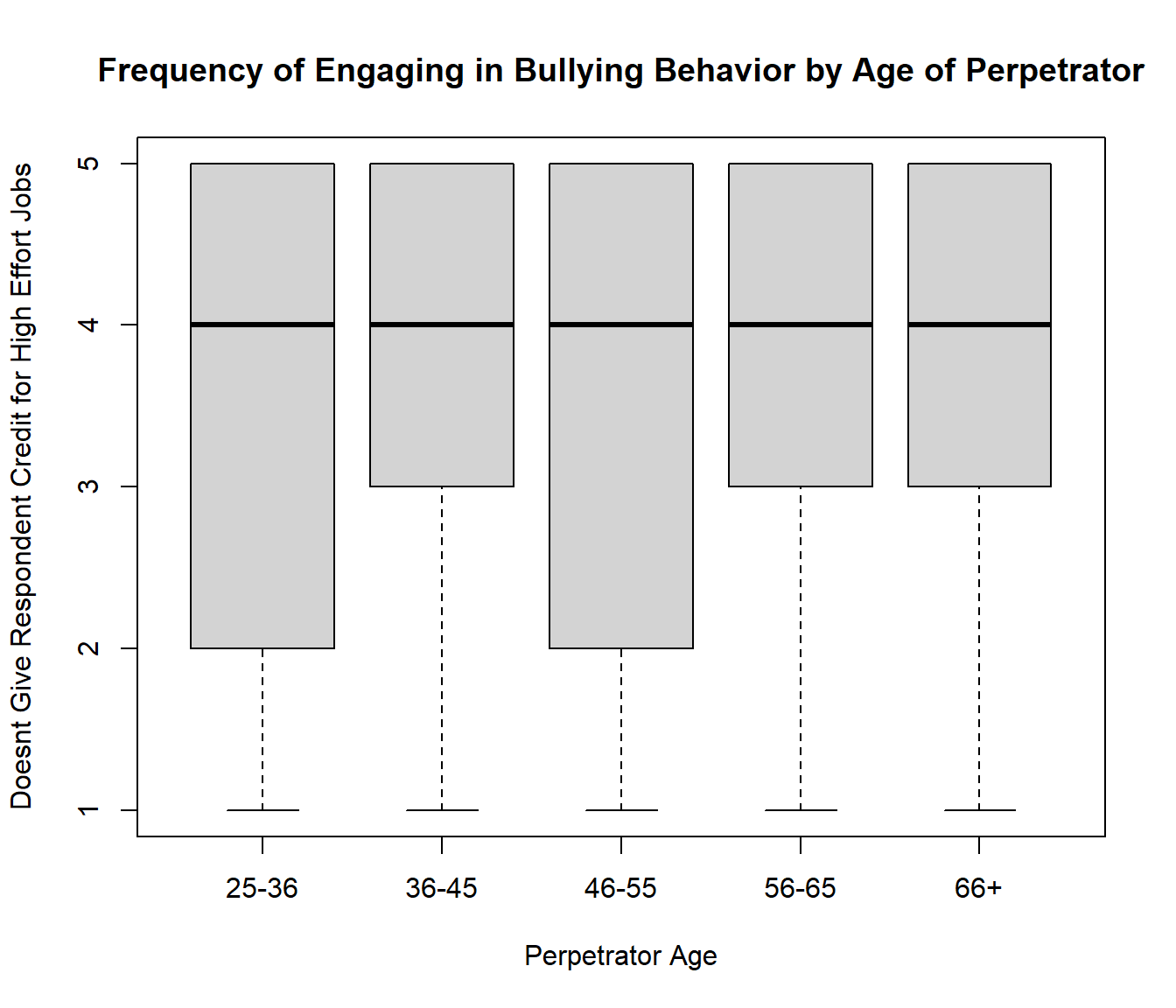

```
##
##  Pearson's Chi-squared test
##
## data:  table12_13
## X-squared = 41.009, df = 16, p-value = 0.0005537
##
##  Pairwise comparisons using Wilcoxon rank sum test with continuity correction
##
## data:  ds$Q12_13 and ds$Q8
##
##   1     2     3     4
## 2 0.112 -     -     -
## 3 0.112 0.880 -     -
## 4 0.011 0.112 0.112 -
## 5 0.112 0.797 0.797 0.662
##
## P value adjustment method: BH
```

**12_14. Breaks Promises**

There is a significant relationship (p<0.1) between the bullying behavior “breaking promises” and perpetrator age groups “25-35” and all age groups except “66+.”

| Frequency of Breaking Promises by Age of Perpetrator | | | | | |
| --- | --- | --- | --- | --- | --- |
|  | **Never** | **Seldom** | **Occasional** | **Moderately Often** | **Very Often** |
| 25-36 | 27 | 4 | 7 | 13 | 24 |
| 36-45 | 88 | 27 | 41 | 78 | 176 |
| 46-55 | 88 | 29 | 63 | 80 | 202 |
| 56-65 | 63 | 35 | 43 | 47 | 140 |
| 66+ | 16 | 7 | 11 | 9 | 29 |

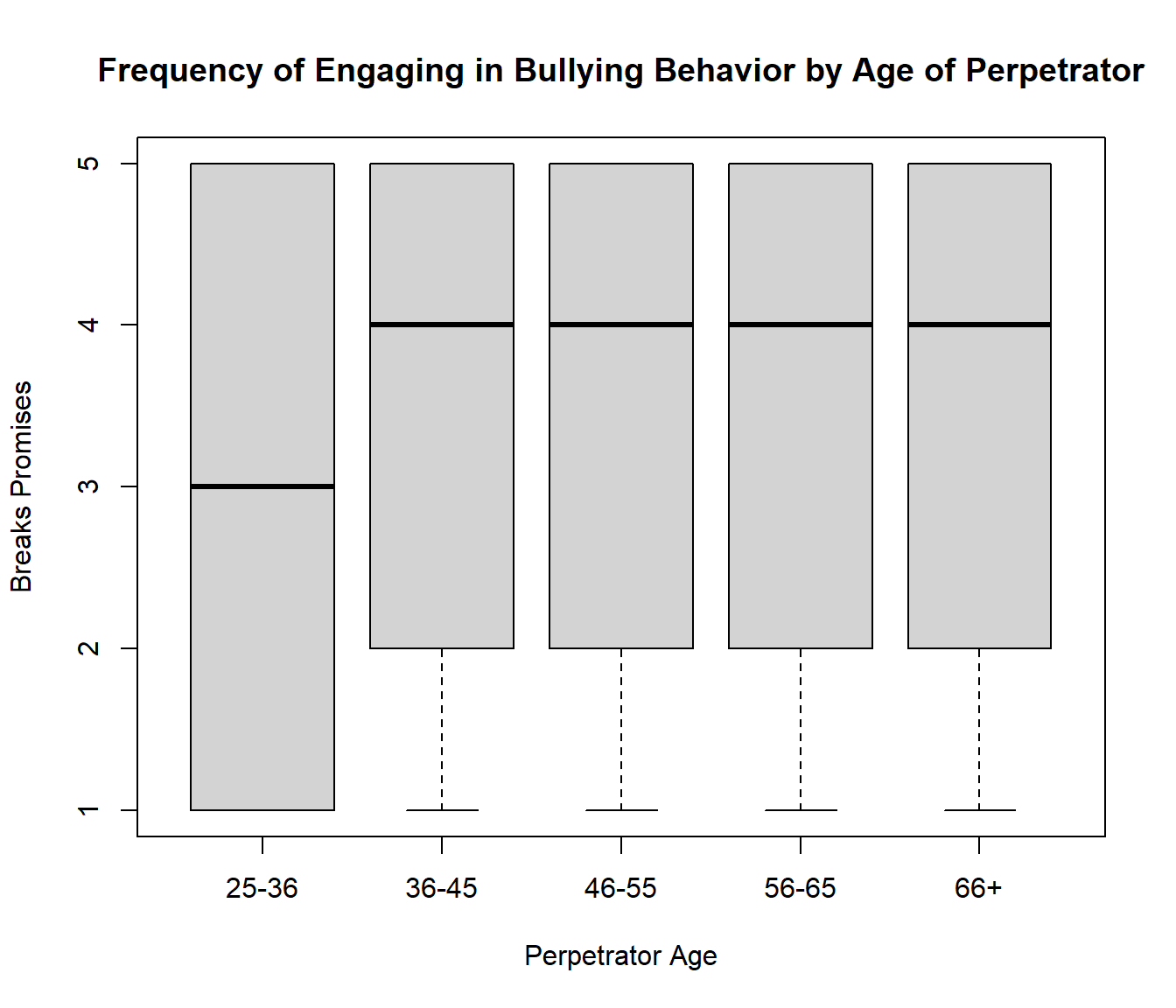

```
##
##  Pearson's Chi-squared test
##
## data:  table12_14
## X-squared = 25.298, df = 16, p-value = 0.06475
##
##  Pairwise comparisons using Wilcoxon rank sum test with continuity correction
##
## data:  ds$Q12_14 and ds$Q8
##
##   1     2     3     4
## 2 0.079 -     -     -
## 3 0.077 0.758 -     -
## 4 0.081 0.758 0.680 -
## 5 0.473 0.680 0.660 0.731
##
## P value adjustment method: BH
```

**12_15. Lying**

| Frequency of Lying to Respondent by Age of Perpetrator | | | | | |
| --- | --- | --- | --- | --- | --- |
|  | **Never** | **Seldom** | **Occasional** | **Moderately Often** | **Very Often** |
| 25-36 | 17 | 13 | 7 | 12 | 27 |
| 36-45 | 82 | 28 | 55 | 74 | 174 |
| 46-55 | 95 | 51 | 57 | 74 | 189 |
| 56-65 | 59 | 36 | 46 | 43 | 146 |
| 66+ | 21 | 3 | 10 | 7 | 30 |

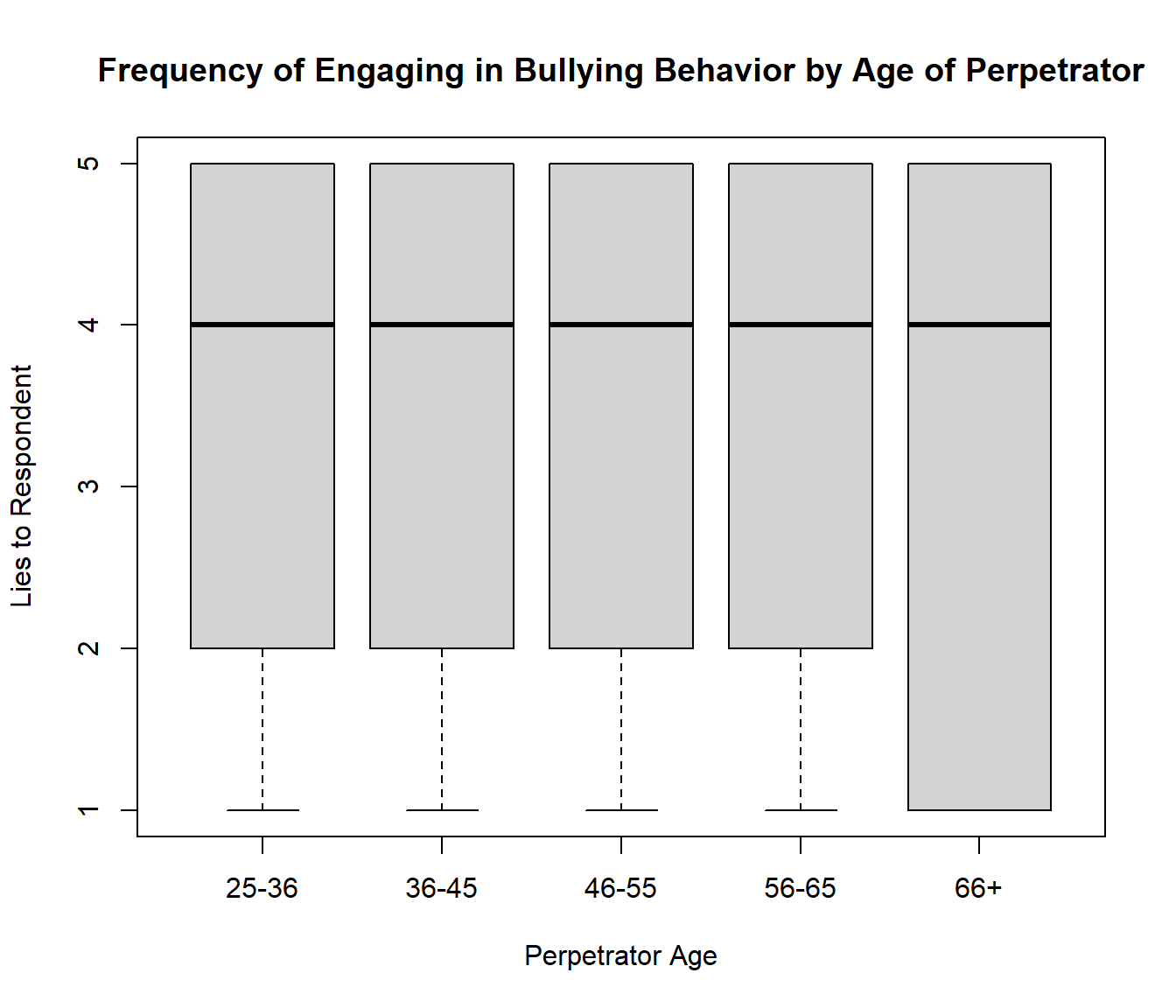

```
##
##  Pearson's Chi-squared test
##
## data:  table12_15
## X-squared = 22.696, df = 16, p-value = 0.1221
```

**Data analysis of the checklist of bullying behaviors with perpetrator age group**

**13_1. Bad Recommendation**

There is a significant relationship (p<0.05) between the bullying behavior “giving a bad recommendation” and perpetrator age groups “25-35” and “46-55”; “25-35” and “56-65”; and “36-45” and “56-65.”

| Frequency of Giving a Bad/Unfair Recommendation by Perpetrator Age | | |
| --- | --- | --- |
| Age group | **Yes** | **No** |
| 25-36 | 49 | 25 |
| 36-45 | 220 | 184 |
| 46-55 | 236 | 227 |
| 56-65 | 142 | 182 |
| 66+ | 37 | 33 |

```
##
##  Pearson's Chi-squared test
##
## data:  table1
## X-squared = 15.527, df = 4, p-value = 0.003724
##
##  Pairwise comparisons using Wilcoxon rank sum test with continuity correction
##
## data:  ds$Q13_1 and ds$Q8
##
##   1      2      3      4
## 2 0.1223 -      -      -
## 3 0.0493 0.3823 -      -
## 4 0.0051 0.0220 0.1213 -
## 5 0.1734 0.8049 0.8049 0.2423
##
## P value adjustment method: BH
```

**13_2. Cancel Visa**

| Frequency of Cancelling or Threatening to Cancel Respondents Visa/Work Permit by Perpetrator Age | | |
| --- | --- | --- |
| Age group | **Yes** | **No** |
| 25-36 | 70 | 3 |
| 36-45 | 366 | 38 |
| 46-55 | 405 | 50 |
| 56-65 | 287 | 36 |
| 66+ | 63 | 7 |

```
##
##  Pearson's Chi-squared test
##
## data:  table2
## X-squared = 3.8803, df = 4, p-value = 0.4224
```

**13_3. Lengthen Stay in Lab**

There is a significant relationship (p<0.05) between the bullying behavior “lengthening stay in lab” and perpetrator age groups “25-35” and all other age groups.

| Frequency of Unnecessarily Lengthening Respondents Stay in his/her Lab by Perpetrator Age | | |
| --- | --- | --- |
| Age group | **Yes** | **No** |
| 25-36 | 61 | 13 |
| 36-45 | 265 | 143 |
| 46-55 | 300 | 156 |
| 56-65 | 212 | 109 |
| 66+ | 45 | 27 |

```
##
##  Pearson's Chi-squared test
##
## data:  table3
## X-squared = 9.4846, df = 4, p-value = 0.05006
##
##  Pairwise comparisons using Wilcoxon rank sum test with continuity correction
##
## data:  ds$Q13_3 and ds$Q8
##
##   1     2     3     4
## 2 0.018 -     -     -
## 3 0.018 0.885 -     -
## 4 0.018 0.885 0.942 -
## 5 0.018 0.885 0.885 0.885
##
## P value adjustment method: BH
```

**13_4. Take Away Funding**

There is a significant relationship (p<0.05) between the bullying behavior “lengthening stay in lab” and perpetrator age groups “25-35” and all other age groups.

| Frequency of Taking Away or Respondents Funding or Threatening to Take Away Funding by Perpetrator Age | | |
| --- | --- | --- |
| Age group | **Yes** | **No** |
| 25-36 | 60 | 15 |
| 36-45 | 241 | 167 |
| 46-55 | 245 | 212 |
| 56-65 | 177 | 142 |
| 66+ | 38 | 34 |

```
##
##  Pearson's Chi-squared test
##
## data:  table4
## X-squared = 19.867, df = 4, p-value = 0.0005305
##
##  Pairwise comparisons using Wilcoxon rank sum test with continuity correction
##
## data:  ds$Q13_4 and ds$Q8
##
##   1       2       3       4
## 2 0.00149 -       -       -
## 3 0.00019 0.21305 -       -
## 4 0.00049 0.47519 0.75268 -
## 5 0.00149 0.47519 0.89572 0.75268
##
## P value adjustment method: BH
```

**13_5. Encourage Others to Mistreat**

There is a significant relationship (p<0.01) between the bullying behavior “encouraging others to mistreat” and perpetrator age groups “36-45” and “56-65”.

| Frequency of Encouraging Others to Mistreat Respondent by Perpetrator Age | | |
| --- | --- | --- |
| Age group | **Yes** | **No** |
| 25-36 | 35 | 40 |
| 36-45 | 205 | 202 |
| 46-55 | 224 | 241 |
| 56-65 | 124 | 204 |
| 66+ | 33 | 39 |

```
##
##  Pearson's Chi-squared test
##
## data:  table5
## X-squared = 12.883, df = 4, p-value = 0.01186
##
##  Pairwise comparisons using Wilcoxon rank sum test with continuity correction
##
## data:  ds$Q13_5 and ds$Q8
##
##   1      2      3      4
## 2 0.7950 -      -      -
## 3 0.8991 0.7950 -      -
## 4 0.5182 0.0067 0.0189 -
## 5 0.9214 0.7950 0.8903 0.5182
##
## P value adjustment method: BH
```

**13_6. Use Data without Acknowledging Contribution**

There is a weak significant relationship (p<0.2) between the bullying behavior “using data without acknowledging contribution” and perpetrator age groups “25-35” and “36-45”; “25-35” and “46-55”; and “36-45” and “56-65”.

| Frequency of Using Respondents Data in Paper/Patents without Acknowledging Contribution by Perpetrator Age | | |
| --- | --- | --- |
| Age group | **Yes** | **No** |
| 25-36 | 55 | 20 |
| 36-45 | 236 | 169 |
| 46-55 | 283 | 175 |
| 56-65 | 213 | 109 |
| 66+ | 48 | 24 |

```
##
##  Pearson's Chi-squared test
##
## data:  table6
## X-squared = 9.305, df = 4, p-value = 0.05391
##
##  Pairwise comparisons using Wilcoxon rank sum test with continuity correction
##
## data:  ds$Q13_6 and ds$Q8
##
##   1    2    3    4
## 2 0.14 -    -    -
## 3 0.18 0.42 -    -
## 4 0.39 0.15 0.39 -
## 5 0.48 0.39 0.48 0.93
##
## P value adjustment method: BH
```

**13_7. Violate Authorship Guidelines**

There is a significant relationship (p<0.05) between the bullying behavior “violating authorship guidelines” and perpetrator age groups “25-35” and “36-45”; and “25-35” and “46-55.”

| Frequency of Violating Authorship Contribution Guidelines by Perpetrator Age | | |
| --- | --- | --- |
| Age group | **Yes** | **No** |
| 25-36 | 55 | 20 |
| 36-45 | 233 | 173 |
| 46-55 | 258 | 202 |
| 56-65 | 197 | 123 |
| 66+ | 43 | 28 |

```
##
##  Pearson's Chi-squared test
##
## data:  table7
## X-squared = 9.3599, df = 4, p-value = 0.05271
##
##  Pairwise comparisons using Wilcoxon rank sum test with continuity correction
##
## data:  ds$Q13_7 and ds$Q8
##
##   1     2     3     4
## 2 0.049 -     -     -
## 3 0.049 0.778 -     -
## 4 0.189 0.427 0.255 -
## 5 0.255 0.773 0.685 0.876
##
## P value adjustment method: BH
```

**13_8. Sign Away Rights**

| Frequency of Forcing Respondent to Sign Away Rights by Perpetrator Age | | |
| --- | --- | --- |
| Age group | **Yes** | **No** |
| 25-36 | 65 | 9 |
| 36-45 | 341 | 56 |
| 46-55 | 376 | 81 |
| 56-65 | 254 | 61 |
| 66+ | 58 | 12 |

```
##
##  Pearson's Chi-squared test
##
## data:  table8
## X-squared = 4.9834, df = 4, p-value = 0.289
```

**13_9. Violate IP Rights**

There is a significant relationship (p<0.1) between the bullying behavior “violating IP rights” and perpetrator age groups “25-35” and “36-45”; and “25-35” and “56-65.”

| Frequency of Violating Respondents IP Rights by Perpetrator Age | | |
| --- | --- | --- |
| Age group | **Yes** | **No** |
| 25-36 | 60 | 14 |
| 36-45 | 268 | 139 |
| 46-55 | 331 | 130 |
| 56-65 | 212 | 106 |
| 66+ | 53 | 18 |

```
##
##  Pearson's Chi-squared test
##
## data:  table9
## X-squared = 10.467, df = 4, p-value = 0.03325
##
##  Pairwise comparisons using Wilcoxon rank sum test with continuity correction
##
## data:  ds$Q13_9 and ds$Q8
##
##   1     2     3     4
## 2 0.078 -     -     -
## 3 0.238 0.195 -     -
## 4 0.078 0.817 0.243 -
## 5 0.442 0.243 0.688 0.275
##
## P value adjustment method: BH
```

**13_10. Cancel Position**

There is a significant relationship (p<0.001) between the bullying behavior “cancelling or threatening to cancel position” and perpetrator age groups “25-35” and all other age groups.

| Frequency of Cancelling or Threatening to Cancel Respondents Current Appointment/Position by Perpetrator Age | | |
| --- | --- | --- |
| Age group | **Yes** | **No** |
| 25-36 | 54 | 20 |
| 36-45 | 189 | 218 |
| 46-55 | 216 | 240 |
| 56-65 | 140 | 187 |
| 66+ | 31 | 42 |

```
##
##  Pearson's Chi-squared test
##
## data:  table10
## X-squared = 23.006, df = 4, p-value = 0.0001263
##
##  Pairwise comparisons using Wilcoxon rank sum test with continuity correction
##
## data:  ds$Q13_10 and ds$Q8
##
##   1       2       3       4
## 2 0.00014 -       -       -
## 3 0.00015 0.87182 -       -
## 4 2.8e-05 0.54485 0.41435 -
## 5 0.00048 0.66420 0.62341 0.95728
##
## P value adjustment method: BH
```

**Visual indication of the trends of all contextual bullying behaviors across various age groups.**

Gave Bad/Unfair Recommendation

| Age Group | Percentage of Perpetrators Who... |
| --- | --- |
| 1 | 33.8 |
| 2 | 45.5 |
| 3 | 48.6 |
| 4 | 56.2 |
| 5 | 47.1 |

Cancelled or Threatened to Cancel My Visa

| Age Group | Percentage of Perpetrators Who… |
| --- | --- |
| 1 | 4.1 |
| 2 | 9.4 |
| 3 | 10.2 |
| 4 | 11.1 |
| 5 | 10 |

Unnecessarily Lengthened My Stay in His/Her Lab

| Age Group | Percentage of Perpetrators Who… |
| --- | --- |
| 1 | 17.6 |
| 2 | 35 |
| 3 | 33.6 |
| 4 | 34 |
| 5 | 37.5 |

Took Away Funding or Threatened to Take Away Funding

| Age Group | Percentage of Perpetrators Who… |
| --- | --- |
| 1 | 20 |
| 2 | 40.9 |
| 3 | 45.8 |
| 4 | 44.5 |
| 5 | 47.2 |

Encouraged Others to Mistreat Me

| Age Group | Percentage of Perpetrators Who… |
| --- | --- |
| 1 | 53.3 |
| 2 | 49.6 |
| 3 | 51.4 |
| 4 | 62.2 |
| 5 | 54.2 |

Used My Data in Papers/Patents without Acknowledging My Contribution

| Age Group | Percentage of Perpetrators Who… |
| --- | --- |
| 1 | 26.7 |
| 2 | 41.7 |
| 3 | 37.7 |
| 4 | 33.9 |
| 5 | 33.3 |

Violated Authorship Contribution Guidelines

| Age Group | Percentage of Perpetrators Who… |
| --- | --- |
| 1 | 26.7 |
| 2 | 42.6 |
| 3 | 43.4 |
| 4 | 38.4 |
| 5 | 39.4 |

Forced Me to Sign Away My Rights

| Age Group | Percentage of Perpetrators Who… |
| --- | --- |
| 1 | 12.2 |
| 2 | 14.1 |
| 3 | 17 |
| 4 | 19.4 |
| 5 | 17.1 |

Violated My Intellectual Property Rights

| Aage Group | Percentage of Perpetrators Who… |
| --- | --- |
| 1 | 18.9 |
| 2 | 34.2 |
| 3 | 27.6 |
| 4 | 33.3 |
| 5 | 25.4 |

Cancelled or Threatened to Cancel My Current Appointment/Position

| Age Group | Percentage of Perpetrators Who… |
| --- | --- |
| 1 | 27 |
| 2 | 53.6 |
| 3 | 52.6 |
| 4 | 57.2 |
| 5 | 57.5 |
